# Immunity to *Streptococcus pyogenes* and Common Respiratory Viruses at Age 0-4 Years after COVID-19 restrictions: A Cross-Sectional Study

**DOI:** 10.1101/2025.04.10.25325549

**Authors:** Kitt Dokal, Samuel Channon-Wells, Catherine Davis, Diego Estrada-Rivadeneyra, Kristin K Huse, Amelia Lias, Shea Hamilton, Rebecca L Guy, Theresa Lamagni, Sam Nichols, Andrew Taylor, Philipp KA Agyeman, Amutha Anpananthar, Romain Basmaci, Enitan D Carrol, Michael J Carter, Tisham De, Marien I de Jonge, Marieke Emonts, Leire Estramiana Elorrieta, Katy Fidler, Mojca Kolnik, Taco W Kuijpers, Federico Martinon-Torres, Henriette Moll, Marine Mommert-Tripon, Samira Neshat, Maggie Nyirenda-Nyang’wa, Sean O’Riordan, Daniel R Owens, Nazima Pathan, Stephane Paulus, Mark J Peters, Marko Pokorn, Andrew J Pollard, Irene Rivero Calle, Pablo Rojo, Lorenza Romani, Prita Rughani, Luregn J Schlapbach, Nina A Schweintzger, Ching-Fen Shen, Artur Sulik, Maria Tsolia, Effua Usuf, Michiel van der Flier, Clementien Vermont, Ulrich von Both, Paul Wellman, Victoria J Wright, Shunmay Yeung, Dace Zavadska, Aubrey J Cunnington, Colin Fink, Jethro Herberg, Myrsini Kaforou, PERFORM Consortium, DIAMONDS Consortium, Shiranee Sriskandan, Michael Levin, Tom Parks

## Abstract

**Importance:** The upsurge in invasive disease caused by *Streptococcus pyogenes* among children reported in several European countries during 2022-2023 has not been fully explained.

**Objective:** To test the hypothesis that changes in circulation of common respiratory pathogens associated with the introduction of non-pharmaceutical interventions (NPIs) during the COVID-19 pandemic altered acquisition of immunity to *S. pyogenes* and common respiratory viruses.

**Design:** Cross-sectional study recruiting before (September 2016 to March 2020) and after (April 2020 to July 2023) introduction of NPIs.

**Setting:** European hospitals in 10 countries.

**Participants:** Children with suspected infection and afebrile controls, aged 0-4 years.

**Main Outcomes:** Molecular detection of bacterial and viral pathogens on throat swabs and age-stratified total serum IgG reactivity to *S. pyogenes* cell wall extract from two strains (measured by ELISA), and respiratory syncytial virus (RSV), five influenza viruses and four common cold coronaviruses and SARS-CoV-2 measured by Mesoscale immunoassay.

**Results:** Throat swabs from 1942 children under 5-years of age were tested for respiratory pathogens (1449 recruited before introduction of NPIs; 493 recruited after). A decrease in detection of *S. pyogenes,* RSV, common cold coronaviruses, and influenza viruses was observed between March 2020 to July 2021, corresponding to the maximal period of NPIs. Antibodies to *S. pyogenes* were measured in 252 children recruited before NPIs and 200 thereafter. Antibodies to viral antigens were measured in 230 before NPIs and 92 thereafter. Total IgG to *S. pyogenes* and RSV was significantly lower in children aged 3-4 years recruited after NPI introduction compared with before (*S. pyogenes emm1*: after, n=67, median 0.13 vs before, n=87, 0.35 IVIG relative-units, p=0.007. RSV: after, n=30, median 49604 vs before, n=76, 141782 mesoscale-units p<0.001). No such differences were observed for children aged 0-2 years, or for individual influenza viruses, common cold coronaviruses or SARS-CoV-2.

**Conclusions and Relevance:** We found a significant reduction in serum antibodies to *S. pyogenes* and RSV in children aged 3-4 years after introduction of NPIs. Equivalent to approximately a year delay in acquisition of immunity, these data provide a biological basis for the 2022-2023 upsurge in severe *S. pyogenes* infections in this age group.

## Background

A multi-country outbreak of severe invasive *Streptococcus pyogenes* (iGAS) infections was reported in the fourth quarter of 2022, with greatest impact on children under 10 years of age. This featured unusually severe presentations including rapidly progressive empyema, and septic shock.^1–3^ In the United Kingdom (UK), where infections normally peak between the first and second quarter,^4^ both iGAS disease and scarlet fever are notifiable. However, a sharp out-of-season increase in scarlet fever notifications in the fourth quarter was matched by a sharp increase in primary care consultations for streptococcal pharyngitis: both preceded a peak in iGAS notifications.^1^ For example, in the UK, surveillance of *S. pyogenes* strains causing iGAS demonstrated an initial expansion of *emm12* strains in the second and third quarters of 2022, followed by a rapid increase in *emm1 S. pyogenes* strains – mirrored in several European countries – in the fourth quarter of 2022, such that *emm1* accounted for approximately 70% of severe paediatric infections.^4–7^ Many of the children admitted to hospital with severe iGAS infection were reported to have coinfections with respiratory viruses, in particular RSV.^4,8^ Moreover, coinfection with respiratory viruses was identified in more than half of the UK children who died in the community from iGAS infection during this period. ^9^

Reduced exposure to common respiratory viruses and bacteria during the period of COVID-19-related restrictions has been postulated to result in loss of population immunity to common childhood pathogens.^10^ Scarlet fever is a disease that most commonly affects children in their first year at school.^11^ In the UK, and other European countries, school attendance was markedly reduced from March 2020 leading to reduced exposure to *S. pyogenes*, rates of which fell markedly during the period March 2020-March 2022.^2,12^ The term “immunity debt” was proposed by Cohen *et al.,* who suggested that reduced circulation of common pathogens as a result of non-pharmaceutical interventions (NPIs) resulted in impaired community immunity to common infectious agents, and thus to an increase in severe childhood illness when normal social interaction returned.^13^ The concept of immune debt (also called “immunity gap”) has been widely debated in the scientific literature and popular press.^14,15^ However, although it is a plausible explanation for the increase in childhood illness following relaxation of pandemic social distancing measures, there is so far little biological data to support this hypothesis.^16^

Accordingly, using serum samples from children presenting to hospital as an indicator of community-level immunity in children, we investigated whether a reduction in acquisition of antibody-mediated immunity to common respiratory pathogens could be detected when comparing samples collected before and after introduction of NPIs in March 2020.

## Methods

### i) Study design

Our study comprised: 1) a cross-sectional pathogen detection study covering September 2016 to July 2023 and 2) a cross-sectional study of antibody-mediated immunity to *S. pyogenes* and common respiratory viruses before and after introduction of NPIs in March 2020. Both investigations focused on children attending hospitals in Europe recruited to one of two EU-funded prospective, multicentre observational studies of febrile illness during childhood: PERFORM^17^ recruiting from 19 hospitals in nine European countries between September 2016 and March 2020, and DIAMONDS recruiting from 20 hospitals in six European countries between March 2020 and July 2023.^18^

Across both studies children aged 0-18 years presenting with fever or suspected infection who were sufficiently unwell to require blood tests were recruited from emergency departments, paediatric inpatient wards and paediatric intensive care units. Additionally non-febrile children who were undergoing blood sampling for unrelated reasons were recruited as controls, providing they had not experienced fever or received a vaccination in the previous three weeks.^17^ As previously described, each child recruited to the study was assigned a final diagnostic category on the basis of the available clinical, laboratory and imaging data by at least two experienced paediatricians using an algorithm modified from previous iterations^19^ to enhance detection of inflammatory disease (Supplementary Figure 1).

For our pathogen detection study, all children recruited aged 0-4 years to PERFORM and DIAMONDS (including controls) were eligible if a throat swab had been taken during recruitment as part of our centralised molecular testing programme, which was performed retrospectively as previously described to generate a comprehensive view of the pathogens present in the study population.^17^ Additionally, as a sensitivity analysis, we repeated our analysis in all children recruited to the studies aged 0-18 years.

For our antibody-mediated immunity study, we focused on a subset of children aged 0-4 years recruited to PERFORM and DIAMONDS that were not in the diagnostic categories representing definite or potentially life-threatening bacterial or viral infections, or definite inflammatory diagnoses (Supplementary Figure 1). Further details are described in the Supplementary Methods.

### ii) Data sources

De-identified clinical and demographic data were obtained from the PERFORM and DIAMONDS research databases derived from standardised case report forms. Children in definite bacterial, definite viral or unknown bacterial or viral diagnostic categories, and 10% of all other categories, were cross-checked for inconsistencies, as previously described.^17^

For our pathogen detection study, dry flocked throat swabs stored in eNAT™ media were tested retrospectively as part of our centralised molecular testing programme on the MAGPIX® system by Luminex Diasorin, using the NxTAG™ Respiratory Pathogen Panel (RPP).^17^ This gave results for 16 viral and three bacterial targets from which, for this analysis, we focused on influenza A (subtypes H1 and H3), influenza B, respiratory syncytial virus (RSV) (subtypes A and B), and common cold coronaviruses (HKU1, NL63, 229E and OC43). Additionally, we used quantitative PCR (qPCR) to detect *S. pyogenes* targeting the gene for DNase B using a previously described method developed at Micropathology Ltd.^17^ Finally, for samples collected from 2020 onwards, we used a reverse transcription qPCR, also developed at Micropathology Ltd, to detect SARS-CoV-2 targeting the N gene as previously described.^20^

To measure antibody-mediated immunity to *S. pyogenes*, we adapted a previously reported enzyme-linked immunosorbent assay (ELISA) to measure absorbance to *S. pyogenes* cell wall extracts^21,22^ of two representative strain types associated with scarlet fever: H305 (*emm1*/M1)^23^ and H690 (*emm12*/M12).^24^ Further details of the assays are described in the Supplementary Methods. Briefly, following optimisation, cell wall was extracted from overnight cultures of H305 and H690, dialysed overnight and concentrated using a centrifugal filter unit. Extracted cell wall was coated on to 96-well plates, washed, blocked and incubated with serum diluted 1:1000 in blocking buffer in duplicate. Detection was using a Horseradish peroxidase-conjugated goat anti-human IgG Fc secondary antibody (Sigma-Aldrich®; Cat. A0170) with 3,3′,5,5′-Tetramethylbenzidine (Sigma-Aldrich®; Cat. T0565). Absorbance readings are reported relative to IVIG (Privigen, CSL Behring).

To measure antibody-mediated immunity to common respiratory viruses, sera from a subset of children diluted 1:5000 was further tested for total IgG reactivity to viral analytes using the MSD V-PLEX Respiratory Panel 1 and the V-PLEX COVID-19 Coronavirus Panel 3 kits according to the manufacturer’s protocol. Each sample was tested for reactivity to 15 analytes from which, for this analysis, we focused on: hemagglutinin from two influenza A viruses (A/Michigan/45/2015[H1N1] and A/Hong Kong/4801/2014[H3N2]) and two influenza B viruses (B/Phuket/3073/2013 and B/Brisbane/60/2008); prefusion F-protein from RSV; spike proteins from four common cold coronaviruses (HKU1, NL63, 229E and OC43); and nucleocapsid, spike and spike receptor binding domain (RBD) from SARS-CoV-2.

Finally, to add context to our findings, we reviewed statutory notifications of iGAS and scarlet fever to the UK Health Security Agency (UKHSA) from across England made between January 2016 and December 2022. Further details are described in the Supplementary Methods.

### iii) Statistical analysis

Data were inspected for missing and outlying points; categorical variables were tabulated, and continuous variables were summarised in histograms. All analyses were performed using R version 4.3.2^25^ and plots were generated using the *ggplot2* package.^26^ Our sample size was determined by the number of samples available at the time that we conducted the experiments and capacity to run samples on the MSD platform rather than a formal sample size calculation.

For the pathogen detection study, data were transformed to a times series using the R package *tsibble*.^27^ Monthly totals and the proportion of positive samples were calculated with both the two influenza A subtypes and the four common cold coronaviruses each pooled together.

For the antibody-mediated immunity study, we divided children into two periods: those recruited before the pandemic, defined as having a serum sample dated before 23^rd^ March 2020, the start of the first government mandated lockdown in the UK (“non-pharmaceutical intervention”, NPI); and those recruited during or after the pandemic, defined as having a serum sample dated after 22^nd^ April 2020. For simplicity, time before 23^rd^ March 2020 is termed ‘before NPI’ while time after 22^nd^ April 2020 is termed ‘after NPI’ with a month gap to reflect at least one half-life of natural IgGs.^28^

To assess the relationship between age and reactivity before and after introduction of NPIs, we plotted absorption to *S. pyogenes* relative to IVIG and reactivity to the viral antigens against age as a continuous variable. For subsequent analyses, we divided age at time of sampling into six bands beginning at: 0 months, 6 months, 1 year, 2 years, 3 years and 4 years. The first year was divided in half to allow appreciation of the decay of maternally derived antibody. We tested for differences relative to pandemic within each age band using a Wilcoxon rank-sum test, or the Kruskal-Wallis in sensitivity analyses with additional time periods.

To understand changes in the incidence of iGAS and scarlet fever among children aged 3-4 years, relative to those aged 0-2 years, we calculated yearly incidence rates for each disease for each age band based on the total number of notifications divided by the UK Office for National Statistics population estimate for England for that each bracket. We then examined the change in incidence of each disease in age band relative to the respective pre-pandemic average for 2016 to 2019, assessing statistical significance using Poisson regression and investigated interactions between age band and year using likelihood-ratio tests. We did not make adjustment for multiple testing because our primary analysis focused on changes in acquisition of immunity to S. pyogenes, and we did not consider it equally likely that all age-groups or individual viruses would be influenced by timing related to the pandemic.

### v) Research ethics and governance

Ethical approval was obtained at Imperial College London as coordinating site for both PERFORM (Reference: 16/LO/1684) and DIAMONDS (Reference: 20/HRA/1714) as well as each participating centre (Supplementary Tables 1 and 2). All children were recruited with informed parental consent, and assent from older children.

## Results

Between September 2016 and July 2023, a total of 5,484 children aged 0-4 years were recruited to the PERFORM and DIAMONDS studies at sites within Europe. Of these, 1,942 (35.4%) had valid throat swab PCR results available, including 1,449 pre-pandemic samples, and 493 after NPI introduction (Table 1). After exclusions, antibody titres to *S. pyogenes emm1* and *emm12* were available from 452 children aged 0-4 years (153, 33.7%, also with a valid throat swab PCR), while viral panel titres were available for 322 children (103, 32.0%, with a throat swab PCR) (Supplementary Figure 2).

**Table 1.** Baseline characteristics of participants.

| Variable | Pathogen detection |  | Antibody mediated immunity |  |  |  |
| --- | --- | --- | --- | --- | --- | --- |
| Assay | Multiplex PCR |  | ELISA to *S. pyogenes* |  | MSD multiplex |  |
| Stage | Pre (n=1449) | Post (n=493) | Pre (n=252) | Post (n=200) | Pre (n=230) | Post (n=92) |
| Male sex, n (%) | 798 (55.1) | 269 (54.7) | 147 (58.3) | 127 (63.5) | 136 (59.1) | 60 (65.2) |
| Age (months), |  |  |  |  |  |  |
| Median (IQR) | 19.7 (8.2-38.1) | 20.7 (9.1-38.1) | 26.8 (11.6-45.3) | 25.3 (12.2-42.8) | 26.3 (11.3-43.0) | 25.7 (15.3-42.2) |
| Ethnicity, n (%) |  |  |  |  |  |  |
| Afro-Caribbean | 6 (0.4) | 0 | <5 | <5 | <5 | <5 |
| Asian | 89 (6.4) | 86 (18.7) | 10 (4.1) | 17 (8.9) | 10 (4.5) | 6 (6.7) |
| Black African | 34 (2.4) | 45 (9.8%) | 6 (2.4) | 19 (9.9) | 5 (2.2) | 5 (5.6) |
| Middle-Eastern | 20 (1.4) | 8 (1.7) | <5 | <5 | <5 | 0 |
| White European | 1155 (82.6) | 272 (59.3) | 218 (89.0) | 128 (67.0) | 197 (88.3) | 65 (73.0) |
| Mixed | 37 (2.6) | 17 (3.7) | <5 | 8 (4.2) | <5 | 5 (5.6) |
| Other | 57 (4.1) | 31 (6.8) | <5 | 16 (8.5) | <5 | 6 (6.7) |
| Country |  |  |  |  |  |  |
| U.K. | 411 (28.4) | 421 (85.4) | 112 (44.4) | 134 (67.0) | 105 (45.6) | 48 (52.2) |
| E.U. | 1038 (71.6) | 72 (14.6) | 140 (55.5) | 66 (33.0) | 125 (54.3) | 44 (47.8) |

| Diagnostic category |  |  |  |  |  |  |
| --- | --- | --- | --- | --- | --- | --- |
| **Definite bacterial** | 183 (12.6) | 55 (11.2) | 0 | 0 | 0 | 0 |
| **Probable bacterial** | 217 (15.0) | 39 (7.9) | 0 | 0 | 0 | 0 |
| **Bacterial syndrome** | 140 (9.7) | 23 (4.7) | 0 | 0 | 0 | 0 |
| **Unknown bacterial or viral** | 217 (15.0) | 50 (10.1) | 0 | 87 (43.5) | 0 | 0 |
| **Viral syndrome** | 122 (8.4) | 65 (13.2) | 0 | 0 | 0 | 0 |
| **Probable viral** | 215 (14.8) | 46 (9.3) | 98 (38.9) | 52 (26.0) | 94 (40.9) | 52 (56.5) |
| **Definite viral** | 165 (11.4) | 108 (22.0) | 0 | 0 | 0 | 0 |
| **Inflammatory syndrome** | 24 (1.7) | 55 (11.1) | 0 | 0 | 0 | 0 |
| **Other phenotype\*** | 71 (4.9) | 47 (9.5) | 0 | 20 (10.0) | 0 | 0 |
| **Control or trivial illness** | 95 (6.6) | 5 (1.0) | 154 (61.1) | 41 (20.5) | 136 (59.1) | 40 (43.5) |

\*Comprising uncertain infection or inflammation, parasitic infections and other unknown cause of illness.

In both the pathogen detection and antibody-mediated immunity subsets, characteristics of the participants were broadly similar before and after introduction of NPIs, although a higher proportion of samples afterwards had non-White European ethnicity and were recruited in the UK. Additionally, unlike before NPIs, assessment of reactivity to *S. pyogenes* after NPIs included 87 children (43.5%) from the unknown bacterial or viral infection category, 19 (9.5%) from uncertain infection or inflammation category, and one in the other cause of illness category.

In the pathogen detection study, a marked decrease in the prevalence of detected pathogens including RSV, common cold coronaviruses, and influenza viruses, was observed from March 2020 to July 2021, corresponding to the periods of social distancing and school closures (Figure 1). For example, RSV subtypes A and B were only detected in 3 of 16 months during the restrictions period while human coronaviruses other than SARS-CoV-2 were only detected in 2 of 16 months. Detection of *S. pyogenes* also declined during that period with the pathogen detected in samples in 5 of 16 months during the restrictions period compared to 36 of 43 of months previously. Similar changes were seen in our wider cohort of children aged 0-18 years (Supplementary Figure 3).

**Figure 1.**
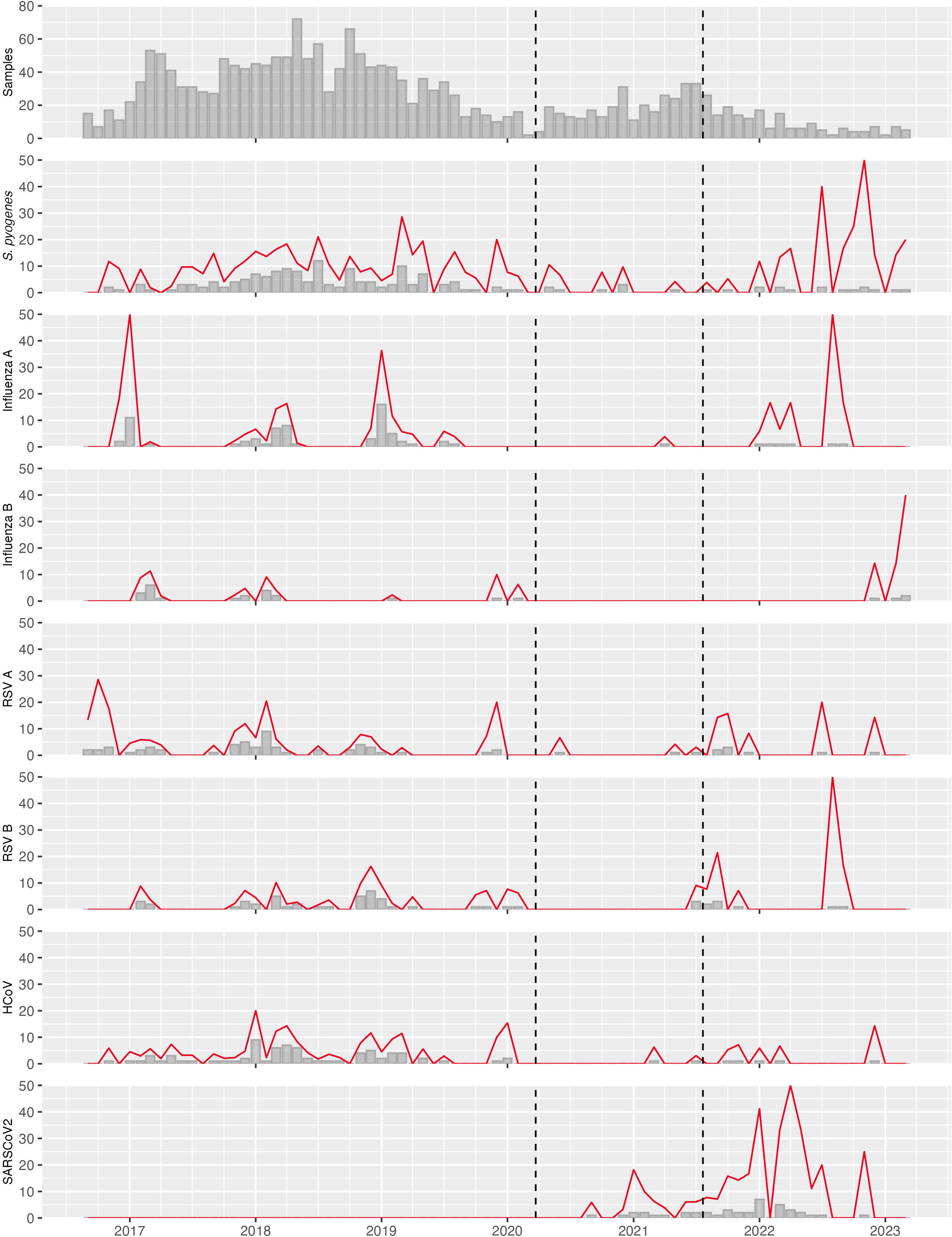
Detection of *S. pyogenes* and viral pathogens by PCR among children aged 0-4 years recruited to the PERFORM and DIAMONDS studies during September 2016 to August 2023. The monthly number of valid samples processed is shown in the first plot followed by the monthly number of positives for each pathogen indicated using grey bars. The red line indicates the monthly percentage of positive samples, plotted on the same scale. Dashed vertical lines indicate the dates of introduction and lifting of restrictions in the UK, as described in the Supplementary Methods. RSV, respiratory syncytial virus; HCoV, common cold coronaviruses 229E, OC43, NL63 and HKU1 (combined); SARSCoV2, Severe acute respiratory syndrome coronavirus 2.

Among 252 children sampled from before the pandemic for the antibody-mediated immunity study, reactivity to *S. pyogenes* cell wall extract from *emm1* and *emm12* – which were highly correlated (*r*=0.94) – declined during the first year of life before rising over the remaining years studied. However, this pattern was altered among the 200 children sampled after introduction of NPIs (Figure 2A and 2B). Excluding 11 children (2.4%) in whom *S. pyogenes* was detectable by PCR at the time of sampling (potentially reflecting acute infection), the median absorbance relative to IVIG among children aged 3-4 years was significantly higher before compared with after (*S. pyogenes*: *emm1* before, n=87, median 0.35 IVIG relative units [RU], interquartile range [IQR], 0.10-0.65 vs *emm1* after, n=67, median, 0.13 RU, IQR 0.04-0.44, p=0.007; *emm12* before, n=87, median 0.51 RU, IQR, 0.18-0.76 vs *emm12* after, n=67, median, 0.21 RU, IQR 0.06-0.59, p=0.005, Figure 3). In multivariate linear regression models, normalised absorbance to both *emm1* and *emm12* was similar among 3-year-old children recruited after NPIs to that of 2-year-old children either before or afterwards (Figure 3A/3B). Similarly, children over 4-years old recruited after the pandemic had similar normalised absorbance to 3-year-old children recruited before pandemic. Further details including sensitivity analyses are described in Supplementary Results.

**Figure 2.**
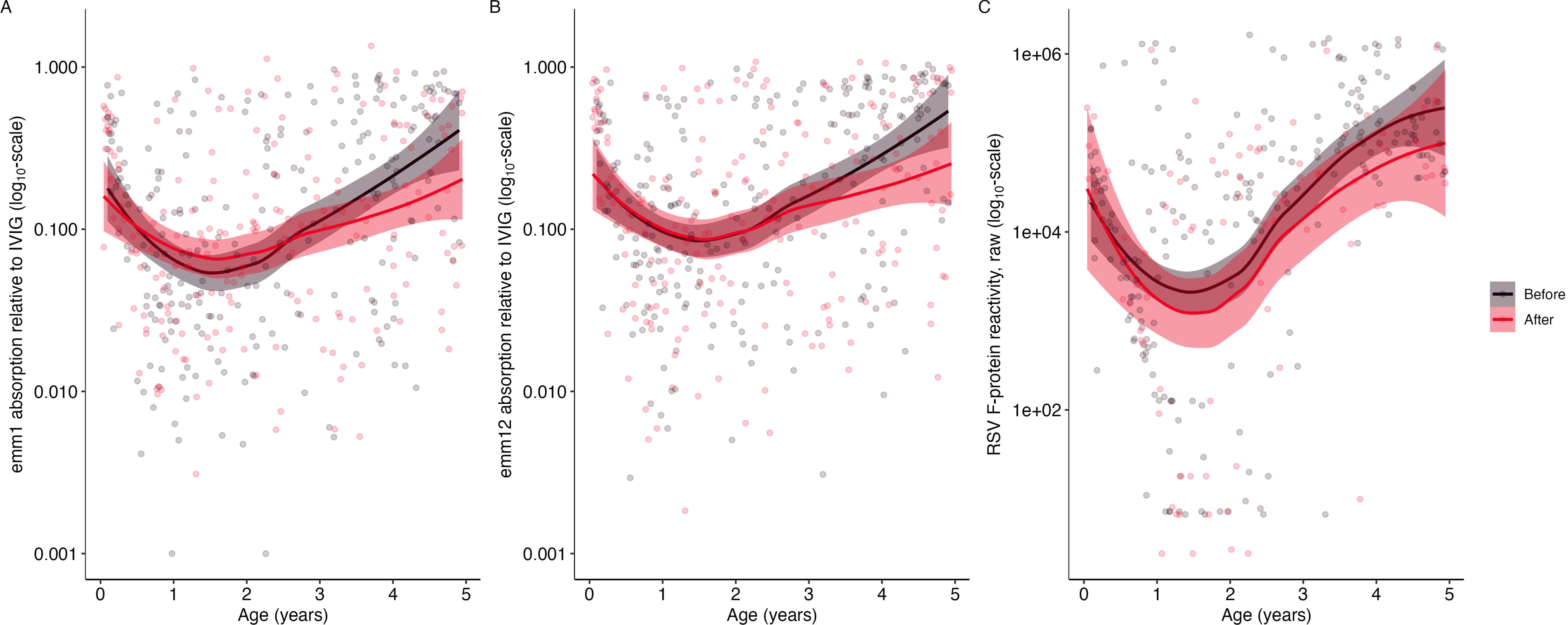
Reactivity to *S. pyogenes* and RSV antigens by age. Scatter plots show reactivity before NPsI (black) and after NPIs (red) March 2020 with a trend line and 95% confidence interval estimated by LOESS regression: **A**. Absorption to M1 relative to IVIG, **B.** Absorption to M12 relative to IVIG, **C.** Reactivity to RSV F-protein.

**Figure 3.**
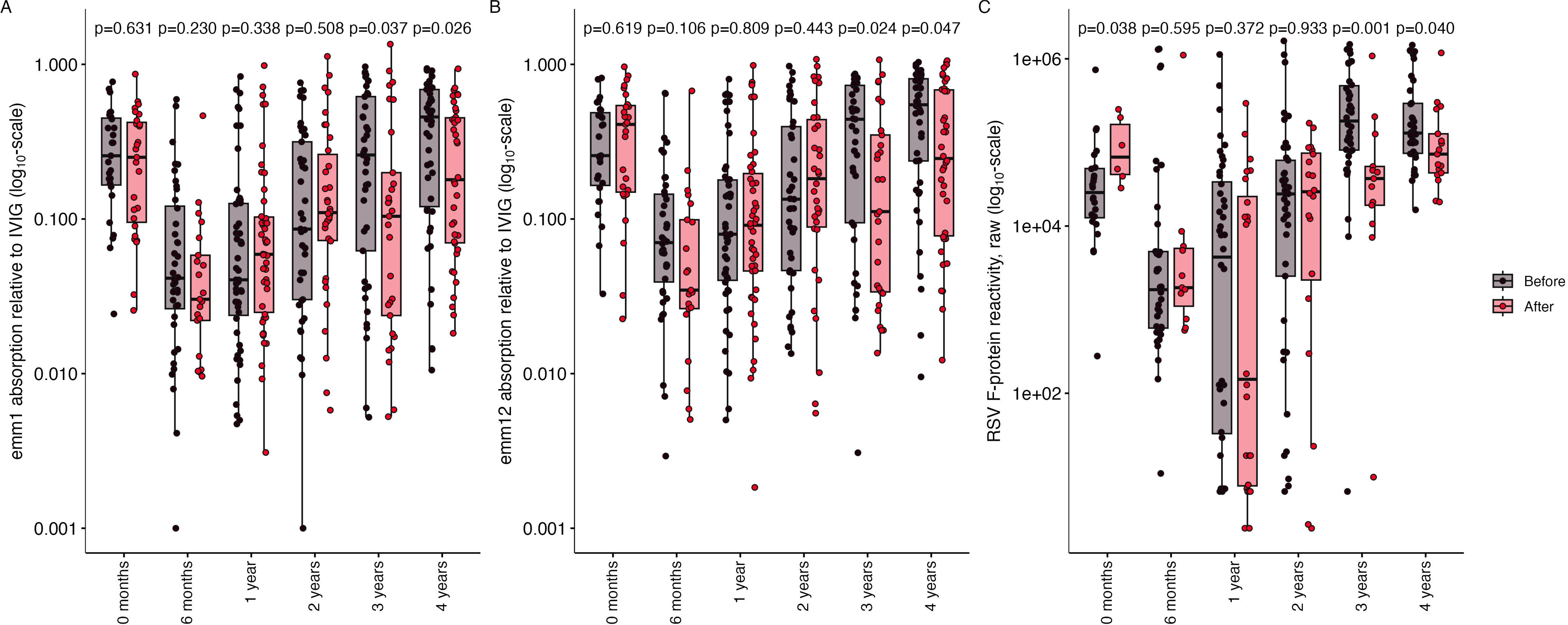
Reactivity to *S. pyogenes* and RSV antigens by age band. Box plots show reactivity before NPIs (black) and after NPIs (red) March 2020 with the difference between groups assessed by a Wilcoxon rank-sum test. **A**. Absorption to *emm1* relative to IVIG, **B.** Absorption to *emm12* relative to IVIG, **C.** Reactivity to RSV F-protein.

A similar age-related acquisition of immunity was seen in a subset of 230 children recruited prior to the pandemic and 92 recruited thereafter in whom antibody reactivity to key antigens from common respiratory viruses were assessed using the MSD assay. Among children aged 3-4 years, excluding eight with detectable RSV at the time of sampling, reactivity to RSV was significantly lower among children recruited after introduction of NPIs (Figure 3C; before, n=76, median 141782 mesoscale units [MU], IQR, 78103-423103, after, n=30, median 49604 MU, IQR, 31065-120655, p<0.001). The statistical relationship between RSV reactivity and age was broadly similar to that observed for *S. pyogenes* although, allowing for the smaller sample size, the effect of NPIs was only statistically significant in the children aged 3 years (Supplementary Table 5).

Correlation between reactivity to individual antigens was mostly explained by age but otherwise weak unless originating from the same or closely related viruses (Supplementary Figure 5). Although some other age-related changes were apparent, there were no consistent pandemic-related patterns of reactivity to individual influenza (Supplementary Figure 6) or common cold coronavirus antigens (Supplementary Figure 7). However, unlike the influenza viruses, reactivity to common cold coronaviruses in aggregate was also lower in children aged 3-4 years recruited after NPIs (Figure 4). In contrast to the other viruses, reactivity to SARS-CoV-2 antigens increased after NPIs, a pattern that was most apparent in samples obtained from July 2021 onwards (Supplementary Figure 8). However, this increase was not apparent in all age groups (Supplementary Figure 9).

**Figure 4.**
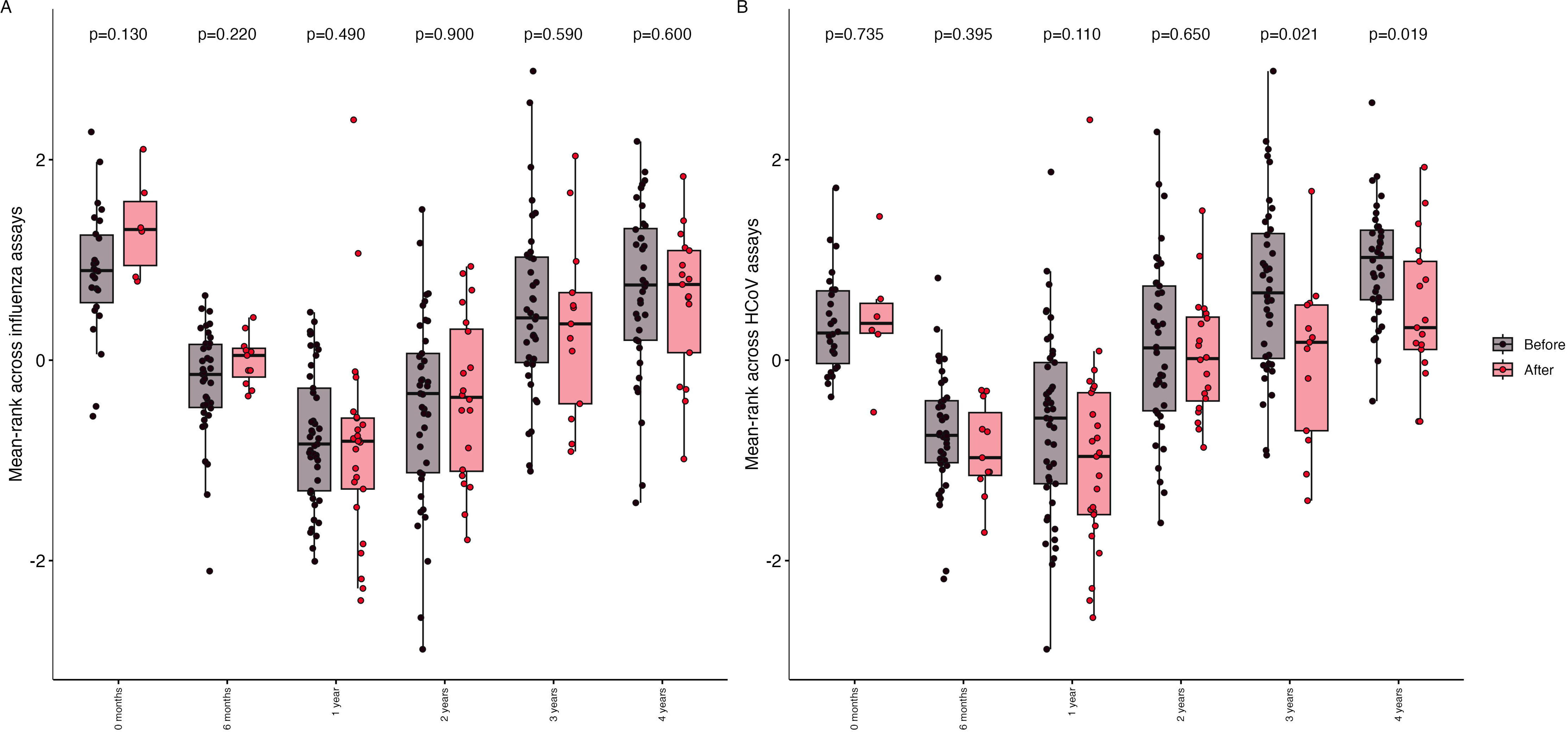
Aggregate reactivity to **A**. circulating influenza A and influenza B viruses and **B**. common cold coronaviruses

Finally, in comparison to the pre-pandemic average, UKHSA notification data for England indicated a significantly greater increase in the incidence rate of iGAS infection but not scarlet fever during 2022 among children aged 3-4 years compared to those aged 0-2 years (Supplementary Figure 10).

## Discussion

We found a significant reduction in antibody concentrations against *S. pyogenes* and RSV in children at 3-4 years of age attending hospitals in the UK and Europe in the first three years after introduction of NPIs to combat the COVID-19 pandemic in March 2020. Although we cannot infer causality from these data, this apparent reduction in antibody-mediated immunity coincides with the exact age group who experienced the greatest increase in life-threatening *S. pyogenes* incidence after NPIs were removed. Although Europe-wide data on *S. pyogenes* incidence were not available, our review of UKHSA data for England confirmed this age group experienced the greatest increase. Moreover, the increase in iGAS cases occurred substantially earlier in the season than is typical, coinciding with a more seasonal but nonetheless equally substantial increase in respiratory viruses, to which our data suggest children were also more susceptible. Ultimately these findings serve as a reminder of the vulnerability of children to *S. pyogenes* infection and the urgent need to develop a vaccine to prevent this disease.

The differences observed in our study appear restricted to pre-school-aged children who are in the process of acquiring adaptive immunity to *S. pyogenes.* Nonetheless, our multivariate analyses suggest that on average the antibody-mediated immunity of children recruited after introduction of NPIs is a year behind that of those recruited before the pandemic. Thus, given the vital importance of antibody in preventing *S. pyogenes* infection, scaled to the population this delayed acquisition of immunity is likely to have contributed to the increase of iGAS infections observed during 2022 to 2023. While the dataset pertaining to viral antigens was smaller, acquisition of immunity to RSV also appeared delayed although the effect appeared limited to 3-4 year-old children. To some extent this difference may reflect an artifact of measuring reactivity to a single RSV antigen rather than the composite of the entire *S. pyogenes* cell wall, but further work would be required to investigate whether reactivity to single *S. pyogenes* antigens is acquired at a faster rate relative to the composite. Our data also suggest NPIs may have altered acquisition of immunity to common cold coronaviruses but not influenza viruses, at least as assessed in this multiplex assay. This result may reflect the complex range of factors that influence population-level immunity to influenza viruses including use of influenza vaccinations.

Similar findings relating to *S. pyogenes* and RSV antibody levels have been reported by one study focusing on adult blood donors sampled in New Zealand.^29^ While limited to adults, who experienced a more marginal increase in iGAS during 2022 to 2023^30,31^ that study benefited from repeated measures from the same individuals and assessed individual *S. pyogenes* antigens rather than our composite cell wall extract approach. Thus the concordance with our results in children is notable and supportive of our conclusions. Other authors have reported waning antibody-mediated immunity to RSV in women of child-bearing age and infants in Canada^32^ and across the age-spectrum in the Netherlands.^33^ Another group working in China observed similar age-specific changes in RSV antibodies among children aged 0-4 years, finding a pronounced decline in children aged 3-4 years during the pandemic.^34^

Strengths of our study include a carefully characterised cohort including well defined diagnostic categories and exclusion of concurrent respiratory infections. Keeping in mind the limited extent to which it is possible to sample healthy children, most are representative of those aged 0-4 years in the general population. Although a wider range of diagnostic categories were included among children recruited during and after the pandemic, adjustment for these differences in our multivariate analyses did not alter our results. Additionally, our study is limited by relatively small sample size and to children aged 0-4 years. Accordingly, our estimates of the differences in antibody levels before and after the pandemic may be imprecise, and further work would be needed to investigate whether the reduction in *S. pyogenes* binding persisted further into childhood. Moreover, we would have ideally extended the analysis of reactivity to viral antigens to a larger number of samples but that was beyond the scope of the current project. In future work we will investigate the extent to which the patterns observed for *S. pyogenes* cell wall antigen would be recapitulated in binding to single antigens. Finally, our work was limited to measurement of total IgG to a limited range of antigens, and we have not yet studied other isoforms or subclasses of antibody, or more unbiased approaches to characterising antibody or other aspects of adaptive immunity to *S. pyogenes*.

In summary, our findings provide an immunological explanation for the upsurge in severe *S. pyogenes* infections affecting younger children and the wider population in 2022-2023. Most importantly, these findings strengthen the case for *S. pyogenes* vaccine development to protect vulnerable groups in the wider population.^35^

## Supporting information

Supplementary Appendix (Consortia members, Supplementary methods and results, Supplementary tables and figures)

## Data Availability

A subset of the data that support the findings of this study will be made available upon acceptance for publication in a peer-reviewed journal. This will include deidentified reactivity data for M1, M12 and RSV assays. Complete raw data will not be publicly available due to privacy concerns and ethics restrictions.

## Acknowledgements

The authors are grateful to all patients and families that contributed towards this study, as well as all members of the DIAMONDS and PERFORM consortia, who are listed in the supplementary appendix. The authors acknowledge additional support from the National Institute for Health and Care Research (NIHR) Biomedical Research Centres at Imperial College London and the University of Newcastle, the NIHR Academic Clinical Lectureship program (MJC), and from the Leducq Foundation [RHD23 and VAC02 (SS and TP)].

## Footnote

This research was funded in part by the Wellcome Trust [222098/Z/20/Z and 215539/Z/19/Z]. For the purpose of open access, the author has applied a CC BY public copyright licence to any Author Accepted Manuscript version arising from this submission.

## Contribution statement

T Parks had full access to all the data in the study and takes responsibility for the integrity of the data and the accuracy of the data analysis.

*Concept and design*: T Parks, S Sriskandan, M Levin, A Cunnington

*Acquisition and interpretation of data:* All authors

*Analysis of the data, including statistical analysis:* T Parks, S Channon-Wells, K Dokal, D Estrada-Rivadeneyra

*Drafting of the manuscript*: T Parks, S Channon-Wells, K Dokal, S Sriskandan, M Levin

*Critical review of the manuscript for important intellectual content:* All authors contributed to critically reviewing the manuscript and approved the final version. Everyone is agreeable to being accountable for all aspects of the work.

## Data Sharing Statement

A subset of the data that support the findings of this study will be made available upon acceptance for publication in a peer-reviewed journal. This will include deidentified reactivity data for M1, M12 and RSV assays as used in our main statistical analysis. Complete raw data will not be publicly available due to privacy concerns and ethics restrictions.

