## Supplementary Appendix (Consortia members, Supplementary methods and results, Supplementary tables and figures) for "Immunity to *Streptococcus pyogenes* and Common Respiratory Viruses at Age 0-4 Years after COVID-19 restrictions: A Cross-Sectional Study"

The members of PERFORM ([www.perform2020.org](http://www.perform2020.org)) are: Michael Levin, Aubrey

Baumard, Evangelos Bellos, Giselle D’Souza, Rachel Galassini, Dominic HabgoodCoote, Shea Hamilton, Clive Hoggart, Sara Hourmat, Heather Jackson, Ian Maconochie, Stephanie Menikou, Naomi Lin, Samuel Nichols, Ruud Nijman, Ivonne Pena Paz, Priyen Shah, Hannah Shailes, Ortensia Vito, Clare Wilson, Amina Abdulla, Ladan Ali, Sarah Darnell, Rikke Jorgensen, Sobia Mustafa, Salina Persand, Molly Stevens, Eunjung Kim, Benjamin Pierce, Katy Fidler, Julia Dudley, Vivien Richmond, Emma Tavliavini, Ching-Fen Shen, Ching-Chuan Liu, Shih-Min Wang, Federico Martinón-Torres, Antonio Salas, Fernando Álvez González, Cristina Balo Farto, Ruth Barral-Arca, María Barreiro Castro, Xabier Bello, Mirian Ben García, Sandra Carnota, Miriam Cebey-López, María José Currás-Tuala, Carlos Durán Suárez, Luisa García Vicente, Alberto Gómez-Carballa, Jose Gómez Rial, Pilar Leboráns Iglesias, Nazareth Martinón-Torres, José María Martinón Sánchez, Belén Mosquera Pérez, Jacobo Pardo-Seco, Lidia Piñeiro Rodríguez, Sara Pischedda, Sara Rey Vázquez, Irene Rivero-Calle, Carmen Rodríguez-Tenreiro, Lorenzo Redondo-Collazo, Miguel Sadiki Ora, Antonio Salas, Sonia Serén Fernández, Cristina Serén Trasorras, Marisol Vilas Iglesias, Dace Zavadska, Anda Balode, Arta Bārzdiņa, Dārta Deksne, Dace Gardovska, Dagne Grāvele, Ilze Grope, Anija Meiere, Ieva Nokalna, Jana Pavāre, Zanda Pučuka, Katrīna Selecka, Aleksandra Sidorova, Dace Svile, Urzula Nora Urbāne, Effua Usuf, Kalifa Bojang, Syed M. A. Zaman, Fatou Secka, Suzanne Anderson, Anna RocaIsatou Sarr, Momodou Saidykhan, Saffiatou Darboe, Samba Ceesay, Umberto D’alessandro, Henrie¨ tte A. Moll, Dorine M. Borensztajn, Nienke N. Hagedoorn, Chantal Tan, Clementien L. Vermont, Joany Zachariasse, W. Dik, Philipp Agyeman, Luregn J. Schlapbach, Christoph Aebi, Verena Wyss, Mariama Usman, Eric Giannoni, Martin Stocker, Klara M. Posfay-Barbe, Ulrich Heininger, Sara Bernhard-Stirnemann, Anita Niederer-Loher, Christian Kahlert, Giancarlo Natalucci, Christa Relly, Thomas Riedel, Christoph Aebi, Christoph Berger, Enitan D. Carrol, Stéphane Paulus, Elizabeth Cocklin, Aakash Khanijau, Rebecca Jennings, Joanne Johnston, Simon Leigh, Karen Newall, Sam Romaine, Maria Tsolia, Irini Eleftheriou, Maria Tambouratzi, Antonis Marmarinos Marietta Xagorari, Kelly Syggelou, Colin Fink, Marie Voice, Leo Calvo-Bado, Werner Zenz, Benno Kohlmaier, Nina A. Schweintzger, Manfred G. Sagmeister, Daniela S. Kohlfürst, Christoph Zurl, Alexander Binder, Susanne Hösele, Manuel Leitner, Lena Pölz, Glorija Rajic, Sebastian Bauchinger, Hinrich Baumgart, Martin Benesch, Astrid Ceolotto, Ernst Eber, Siegfried Gallistl, Gunther Gores, Harald Haidl, Almuthe Hauer, Christa Hude, Markus Keldorfer, Larissa Krenn, Heidemarie Pilch, Andreas Pfleger, Klaus Pfurtscheller, Gudrun Nordberg, Tobias Niedrist, Siegfried Rödl, Andrea Skrabl-Baumgartner, Matthias Sperl, Laura Stampfer, Volker Strenger, Holger Till, Andreas Trobisch, Sabine Löffler, Shunmay Yeung, Juan Emmanuel Dewez, Martin Hibberd, David Bath, Alec Miners, Ruud Nijman, Catherine Wedderburn, Anne Meierford, Baptiste Leurent, Ronald de Groot, Michiel Van der Flier, Marien I. de Jonge, Koen van Aerde, Wynand Alkema, Bryan van den Broek, Jolein Gloerich, Alain J. van Gool, Stefanie Henriet, Martijn Huijnen, Ria Philipsen, Esther Willems, G.P.J.M. Gerrits, M. van Leur, J. Heidema, L. de Haan, C.J. Miedema, C. Neeleman, C.C. Obihara, G.A. Tramper-Stranders, Andrew J. Pollard, Rama Kandasamy, Michael J. Carter, Daniel O’Connor, Sagida Bibi, Dominic F. Kelly, Meeru Gurung, Stephen Thorson, Imran Ansari, David R. Murdoch, Shrijana Shrestha, Zoe Oliver, Marieke Emonts, Emma Lim, Lucille Valentine, Karen Allen, Kathryn Bell, Adora Chan, Stephen Crulley, Kirsty Devine, Daniel Fabian, Sharon King, Paul McAlinden, Sam McDonald, Anne McDonnell, Ailsa Pickering, Evelyn Thomson, Amanda Wood, Diane Wallia, Phil Woodsford, Frances Baxter, Ashley Bell, Mathew Rhodes, Rachel Agbeko, Christine Mackerness, Bryan Baas, Lieke Kloosterhuis, Wilma Oosthoek, Tasnim Arif, Joshua Bennet, Kalvin Collings, Ilona van der Giessen, Alex Martin, Aqeela Rashid, Emily Rowlands, Gabriella de Vries, Fabian van der Velden, Joshua Soon, Ulrich von Both, Laura Kolberg, Manuela Zwerenz, Judith Buschbeck, Christoph Bidlingmaier, Vera Binder, Katharina Danhauser, Nikolaus Haas, Matthias Griese, Tobias Feuchtinger, Julia Keil, Matthias Kappler, Eberhard Lurz, Georg Muench, Karl Reiter, Carola Schoen, François Mallet, Karen Brengel-Pesce, Alexandre Pachot, Marine Mommert, Marko Pokorn, Mojca Kolnik, Katarina Vincek, Tina Plankar Srovin, Natalija Bahovec, Petra Prunk, Veronika Osterman, Tanja Avramoska, Taco Kuijpers, Ilse Jongerius, J.M. van den Berg, D. Schonenberg, A.M. Barendregt, D. Pajkrt, M. van der Kuip, A.M. van Furth, Evelien Sprenkeler, Judith Zandstra, G. van Mierlo, and J. Geissler.

The members of DIAMONDS ([www.diamonds2020.eu](http://www.diamonds2020.eu)) are: Michael Levin, Aubrey Cunnington, Jethro Herberg, Myrsini Kaforou, Victoria J. Wright, Evangelos Bellos, Claire Broderick, Samuel Channon-Wells, Samantha Cooray, Tisham De, Giselle D’Souza, Amedine Duret, Ankita Duseja, Leire Estramiana Elorrieta, Diego Estrada-Rivadeneyra, Rachel Galassini, Dominic Habgood-Coote, Shea Hamilton, Heather Jackson, James Kavanagh, Ilana Keren, Mahdi Moradi Marjaneh, Stephanie Menikou, Samuel Nichols, Ruud Nijman, Harsita Patel, Ivana Pennisi, Oliver Powell, Ruth Reid, Priyen Shah, Ortensia Vito, Elizabeth Whittaker, Clare Wilson, Rebecca Womersley, Amina Abdulla, Sarah Darnell, Sobia Mustafa, Pantelis Georgiou, Jesus Rodriguez-Manzano, Nicolas Moser, Ivana Pennisi, Michael Carter, Paul Wellman,Shane Tibby, Jonathan Cohen, Francesca Davis, Julia Kenny,Marie White, Matthew Fish, Aislinn Jennings, Manu Shankar-Hari, Katy Fidler, Dan Agranoff, Vivien Richmond, Mathhew Seal, Saul Faust, Dan Owen, Ruth Ensom, Sarah McKay, Diana Mondo, Mariya Shaji, Rachel Schranz, Prita Rughani, Amutha Anpananthar, Susan Liebeschuetz, Anna Riddell, Divya Divakaran, Louise Han, Nosheen Khalid, Ivone Lancoma-Malcolm, Jessica Schofield, Teresa Simagan, Mark Peters, Alasdair Bamford, Lauran O’Neill, Nazima Pathan, Esther Daubney, Deborah White, Melissa Heightman, Sarah Eisen, Terry Segal, Lucy Wellings, Simon B Drysdale, Nicole Branch, Lisa Hamzah, Heather Jarman, Maggie Nyirenda, Lisa Capozzi, Emma Gardiner, Robert Moots, Magda Nasher, Anita Hanson, Michelle Linforth, Sean O’Riordan, Donna Ellis, Akash Deep, Ivan Caro, Fiona Shackley, Arianna Bellini, Stuart Gormley, Samira Neshat, Barnaby J Scholefield, Ceri Robbins, Helen Winmill, Stéphane C. Paulus, Andrew J. Pollard, Mark Anthony, Sarah Hopton, Danielle Miller, Zoe Oliver, Sally Beer, Bryony Ward, Shrijana Shrestha, Meeru Gurung, Puja Amatya, Bhishma Pokhrel, Sanjeev Man Bijukchhe, Madhav Chandra Gautam, Sarah Kelly, Peter O’Reilly, Sonu Shrestha, Federico Martinón-Torres, Antonio Salas, Fernando Álvez González, Sonia Ares Gómez, Xabier Bello, Mirian Ben García, Fernando Caamaño Viña, Sandra Carnota, María José Curras-Tuala, Ana Dacosta Urbieta, Carlos Durán Suárez, Isabel Ferreiros Vidal, Luisa García Vicente, Alberto Gómez-Carballa, Jose Gómez Rial, Pilar Leboráns Iglesias, Narmeen Mallah, Nazareth Martinón-Torres, José María Martinón Sánchez, Belén Mosquera Pérez, Jacobo Pardo-Seco, Sara Pischedda, Sara Rey Vázquez, Irene Rivero Calle, Carmen Rodríguez-Tenreiro, Lorenzo Redondo-Collazo, Antonio Salas, Sonia Serén Fernández, Marisol Vilas Iglesias, Enitan D Carrol, Elizabeth Cocklin, Rebecca Beckley, Abbey Bracken, Ceri Evans, Aakash Khanijau, Rebecca Lenihan, Nadia Lewis-Burke, Karen Newall, Sam Romaine, Jennifer Whitbread, Maria Tsolia, Irini Eleftheriou, Nikos Spyridis, Maria Tambouratzi, Despoina Maritsi, Antonios Marmarinos, Marietta Xagorari, Lourida Panagiota, Pefanis Aggelos, Akinosoglou Karolina, Gogos Charalambos, Maragos Markos, Voulgarelis Michalis, Stergiou Ioanna, Marieke Emonts, Emma Lim, John Isaacs, Kathryn Bell, Stephen Crulley, Daniel Fabian, Evelyn Thomson, Diane Wallia, Caroline Miller, Ashley Bell, Fabian J.S. van der Velden, Geoff Shenton, Ashley Price, Owen Treloar, Daisy Thomas, Pablo Rojo, Cristina Epalza, Serena Villaverde, Sonia Márquez, Manuel Gijón, Romina Varchetta, Fátima Machín, Laura Cabello, Irene Hernández, Lourdes Gutiérrez, Ángela Manzanares, Taco Kuijpers, Martijn van de Kuip, A.M. van Furth, J.M. van den Berg, Giske Biesbroek, Floris Verkuil, Carlijn W van der Zee, Dasja Pajkrt, Michael Boele van Hensbroek, Dieneke Schonenberg, Mariken Gruppen, Sietse Nagelkerke, Machiel H Jansen, Ines Goetschalckx, Lorenza Romani, Maia De Luca, Sara Chiurchiù, Costanza Tripiciano, Stefania Mercadante, Clementien L. Vermont, Henriëtte A. Moll, Dorine M. Borensztajn, Nienke N. Hagedoorn, Chantal Tan, Joany Zachariasse, W Dik, Ching-Fen Shen, Dace Zavadska, Sniedze Laivacuma, Aleksandra Rudzate, Diana Stoldere, Arta Barzdina, Elza Barzdina, Sniedze Laivacuma, Monta Madelane, Dagne Gravele, Dace Svile, Romain Basmaci, Noémie Lachaume, Pauline Bories, Raja Ben Tkhayat, Laura Chériaux, Juraté Davoust, Kim-Thanh Ong, Marie Cotillon, Thibault de Groc, Sébastien Le, Nathalie Vergnault, Hélène Sée, Laure Cohen, Alice de Tugny, Nevena Danekova, Marine Mommert-Tripon, Karen Brengel-Pesce, Marko Pokorn, Mojca Kolnik, Tadej Avčin, Tanja Avramoska, Natalija Bahovec, Petra Bogovič, Lidija Kitanovski, Mirijam Nahtigal, Lea Papst, Tina Plankar Srovin, Franc Strle, Katarina Vincek, Michiel van der Flier, Wim J.E. Tissing, Roelie M. Wösten-van Asperen, Sebastiaan J Vastert, Daniel C Vijlbrief, Louis J. Bont, Coco R. Beudeker, Philipp Agyeman, Christoph Aebi, Nina Schöbi, Mariama Usman, Stefanie Schlüchter, Luregn Schlapbach, Cornelia Hagmann, Florian Zapf, Philipp Baumann, Barbara Brotschi, Elisa Zimmermann, Marion Meier, Kathrin Weber, Colin Fink, Marie Voice, Leo Calvo-Bado, Michael Steele, Jennifer Holden, Andrew Taylor, Ronan Calvez, Catherine Davies, Benjamin Evans, Jake Stevens, Peter Matthews, Kyle Billing, Werner Zenz, Alexander Binder, Benno Kohlmaier, Daniela S. Kohlfürst, Nina A. Schweintzger, Christoph Zurl, Susanne Hösele, Piyush G. Gampawar, Barbara Kapo, Manuel Leitner, Lena Pölz, Alexandra Rusu, Glorija Rajic, Bianca Stoiser, Martina Strempfl, Manfred G. Sagmeister, Sebastian Bauchinger, Martin Benesch, Astrid Ceolotto, Ernst Eber, Siegfried Gallistl, Harald Haidl, Almuthe Hauer, Christa Hude, Andreas Kapper, Markus Keldorfer, Sabine Löffler, Tobias Niedrist, Heidemarie Pilch, Andreas Pfleger, Klaus Pfurtscheller, Siegfried Rödl, Andrea Skrabl-Baumgartner, Volker Strenger, Elmar Wallner, Maike K. Tauchert, Ulrich von Both, Laura Kolberg, Patricia Schmied, Ioanna Mavridi, Irene Alba-Alejandre, Katharina Danhauser, Nikolaus Haas, Florian Hoffmann, Matthias Griese, Tobias Feuchtinger, Sabrina Juranek, Matthias Kappler, Eberhard Lurz, Esther Maier, Karl Reiter, Carola Schoen, Sebastian Schroepf, Shunmay Yeung, Manuel Dewez, David Bath, Elizabeth Fitchett, Fiona Cresswell, Effua Usuf, Kalifa Bojang, Anna Roca, Isatou Sarr, Momodou Saidykhan, Ebrahim Ndure, Pedro Madrigal, Silvie Fexova, Artur Sulik, Kacper Toczylowski, Dawid Lewandowski.

**Supplementary Methods**

1. **Study design – Selection of participants**

For our antibody mediated immunity study, we aimed to obtain a sample that was as representative as possible of children in the wider population, with the caveat that, due to the smaller number of samples available from March 2020 onwards, it was necessary to include children from a wider range of diagnostic categories. Thus, for the period before introduction of NPIs, we selected children from three diagnostic categories (controls, probable viral infection and trivial illness), whereas, for children recruited during or after the pandemic, we included all children with an aliquot of serum available at the time of the experiments from six diagnostic categories (controls, probable viral infection, trivial illness, uncertain infection or inflammation, unknown bacterial of viral infection, other cause of illness). To increase parity between the groups, we selected children for the pre-pandemic period with an aliquot of serum available if they resembled a child included during or after the pandemic in terms of at least one of age band, season at time of sampling, and site of recruitment. Subsequently, children were excluded if: they had been assessed as immunocompromised; they had received intravenous immunoglobulin (IVIG) prior to sampling; a sample from a previous timepoint had been included; they were recruited during the first month of non-pharmaceutical interventions (NPIs) in the UK (23rd March to 22nd April 2020); or a subsequent review of the case report form revealed age greater than 5 years or lead to a change to their diagnostic category (Supplementary Figure 2). Finally, due to limited capacity on the Meso Scale Discovery (MSD) platform, we limited assessment of antibody-mediated immunity to common respiratory viruses to children in the three overlapping diagnostic categories (i.e. controls, probable viral infection and trivial illness) and randomly removed a further 23 children selected during “shortlisting ” to achieve approximately equally sized age categories among the pre-pandemic samples.

2. **Data sources – Optimisation of *S. pyogenes* cell wall ELISA**

Colonies for H305 and H690 were streaked onto Columbia blood agar, before they were inoculated into 50ml Todd-Hewitt broth and incubated overnight at 37C with 5% CO2. Overnight cultures were centrifuged at 12,000 x g and bacterial pellets were resuspended in 1 ml cell wall extraction buffer (10mM Tris-HCL; 30% w/v raffinose [Sigma-Aldrich®; Cat. R0250]; 0.1kU/ml Mutanolysin [Sigma-Aldrich®; Cat. M9901]; 1mg/ml lysozyme [Sigma-Aldrich®; Cat. L6876]; and 10ul protease inhibitor cocktail III [VWR, cat. no. 535140]). Following incubation for three hours at 37°C,  samples were centrifuged at 15,000 x g for 10 minutes. The supernatant (cell wall extract) was collection, clarified by filtration (0.2µm) and dialysed overnight into PBS solution at 4°C using a 20kDa MWCO cassette (Thermo Scientific™; Cat. 88528). Cell wall extracts were then concentrated using a centrifugal filter unit (Thermo Scientific™; Cat. G9023). Protein concentration for each of the cell wall extracts were measured using a Pierce™ BCA Protein assay kit (Thermo™; Cat 23227) and followed as per the protocol.

Following optimisation in serum samples from healthy adults with non-group A streptococcus infections (without immunosuppression or prior treatment with intravenous immunoglobulin), 1 ug of cell wall extract (H305 and H690) was coated onto the wells of high binding 96-well plates overnight at 4°C. Wash buffer (0.05% [v/v] Tween-20 in phosphate buffered saline [PBS]) was used to wash the plates before 50 µl blocking buffer (5% bovine serum albumin, 0.1% Tween-20, 0.1% normal goat serum [Sigma-Aldrich®; Cat. G9023] in PBS) was added to each well and incubated for 1 hour at room temperature. After a further washing step, 50µl of serum samples (diluted to 1:1000 in blocking buffer) was added in duplicate for 1 hour at room temperature. After washing the plate three times, 50µl of Fc region specific, Horseradish peroxidase-conjugated goat anti-human antibody diluted 1:60,000 in blocking buffer was added and incubated for 1 hour at room temperature. Wells were washed three times before adding 50µl of 3,3′,5,5′-Tetramethylbenzidine (Sigma-Aldrich®; Cat. T0565) for 25 minutes, protected from light. Next, 25µl of 1M sulphuric acid was added and absorbance was immediately measured at 450nm (Thermo Scientific™ Multiskan™ FC Microplate Photometer; Cat. 51119000) and readings were subtracted from a reference wavelength of 570nm. After subtraction of the blank readings, duplicate absorbance readings were averaged. Absorbance readings were recorded relative to 1:1000 IVIG (Privigen, CSL Behring) run in duplicate for each respective plate. Finally, to assess for non-specific binding, adult samples were also tested with blocking solution containing normal rabbit serum, alongside normal rabbit serum incubated instead of patient samples.

For final analysis, two individuals with M1 reactivity below the limit of detection were assigned arbitrary values of 0.001.

3. **Data sources – UKHSA Notification Data**

We reviewed statutory notifications of iGAS and scarlet fever to the UK Health Security Agency (UKHSA) from across England for children aged 0-4 years made between January 2016 December 2022. More specifically, iGAS infection (sterile-site isolates) reports were extracted from routine and reference laboratory national databases, then merged and grouped into 14-day non-repeating episodes.^1^ Scarlet fever notifications between were extracted from the UKHSA notifications of infectious diseases database.^2^

4. **Statistical analysis – Time periods relative to pandemic**

Our main analysis used a division into two time periods as described in the main text. As a sensitivity analysis, we divided the time after 22nd April 2020 into the periods during which the majority of NPIs were in place (termed ‘During’) up to 19th July 2021 and after this date when they were lifted (termed ‘Easing’). This date was chosen because it was when the UK Government moved to ‘step 4’ of its COVID-19 roadmap, including lifting of all remaining limits on social contact.^3^ Notably, as these dates all reflected general trends in use of NPIs across Europe, time periods were applied across the entire dataset, rather than using different dates for each of the regions that recruited to the study.

5. **Statistical analysis – Multiplex data and multivariate analysis**

We combined reactivity to multiple antigens for groups of related viruses (e.g. influenza viruses) by ranking raw MSD values for each individual assay, and then taking the mean rank across all assays for each patient. For multivariate analyses, we transformed absorption to S. pyogenes relative to IVIG and reactivity to the viral antigens to approximate a normal distribution using a rank-based inverse normal transform, using the RankNorm function from the RNOmni package.^4^ We then performed linear regression including covariates selected *a priori* to minimise confounding due to diagnostic category, recruitment site, and biological sex. We also investigated interactions between age band and timing relative to the pandemic using likelihood-ratio tests. Finally, we assessed correlation between age-adjusted reactivity to distinct antigens by calculating the Pearson’s correlation coefficient (r) for each relationship using the corrplot package,^5^ using the residuals from a linear regression model for the rank-normalised reactivity with parameters for each of age bands to adjust for age.

**Supplementary Results**

1. **Multivariate linear regression modelling**

In our analysis of reactivity to *S. pyogenes* cell wall extract, we used multivariate linear regression models to investigate the relationship between age and reactivity. Importantly, the lower absorbance to both *emm1* and *emm12* among children aged 3-4 and 4-5 years recruited during and after the pandemic compared to those recruited beforehand – reflecting a statistically significant interaction between these age bands and the timing of their recruitment – was unaffected by adjustment for sex, diagnostic category or recruitment inside or outside of the UK (Supplementary Table 3 and Supplementary Table 4).

In addition, these patterns were also apparent for *emm1* and *emm12* in sensitivity analyses encompassing inclusion of the 11 children with detection of S. pyogenes by PCR at the time of the serum sample (Supplementary Figure 4A and 4B), exclusion of the three with detection of SARS-CoV-2 (Supplementary Figure 4C and 4D), and dividing the timing of recruitment into three periods separated by the UK introduction of NPIs and easing from July 2021 onwards (Supplementary Figure 4E and 4F). There was also no evidence of statistically significant differences between the batches in which the assays were performed.

**Supplementary Table 1:** Participating clinical recruitment sites for PERFORM

| **Country** | **Participating site** | **Ethical approval number** |
| --- | --- | --- |
| United Kingdom | - St Mary’s Hospital, Imperial College Healthcare NHS Trust, London - Alder Hey Children’s Hospital, Liverpool - Great North Children’s Hospital, Newcastle upon Tyne - John Radcliffe Hospital, Oxford - Royal Alexandra Children’s Hospital, Brighton | 16/LO/1684 |
| Austria | - Medizinische Universität Graz, Graz | 28-518 ex 15/16 |
| Germany | - Dr. von Hauner Children's Hospital, Ludwig- Maximilians-University, Munich | 699-16 |
| Greece | - P. and A. Kyriakou Children’s Hospital, Athens | 415/13.06.16 |
| Latvia | - Children’s Clinical University Hospital, Riga | 1/16-07-14 |
| Netherlands | - Sophia’s Children’s Hospital, Rotterdam - Academic University Medical Center, Amsterdam - Radboud University Medical Center, Nijmegen | NL58103.091.16 |
| Slovenia | - University Medical Centre Ljubljana | 0120-483/2016-3 |
| Spain | - Hospital Clínico Universitario de Santiago de Compostela | 2016/331 |
| Switzerland | - University Children’s Hospital, Universität Bern, Bern | 2016-01835 |

**Supplementary Table 2:** Participating clinical recruitment sites for DIAMONDS

| **Country** | **Participating site** | **Ethical approval number** |
| --- | --- | --- |
| United Kingdom | - St Mary’s Hospital, Imperial College Healthcare NHS Trust, London - Alder Hey Children’s Hospital, Liverpool - Great North Children’s Hospital, Newcastle upon Tyne - Southampton Children’s Hospital, Southampton - Evelina London Children’s Healthcare, London - Royal Alexandra Children's Hospital, Brighton - John Radcliffe Hospital, Oxford - The Leeds Teaching Hospitals NHS Foundation Trust, Leeds - Leicester Children's Hospital, Leicester - Addenbrookes Hospital, Cambridge - University Hospital Lewisham, London - Royal London Hospital and Newham Hospital, Barts Health NHS Trust, London - Great Ormond Street Hospital, London | 20/HRA/1714 |
| Austria | - Medizinische Universität Graz, Graz | 32-401 Ex 19/20 |
| Germany | - Dr. von Hauner Children's Hospital, Ludwig- Maximilians-University, Munich | 20-0568 |
| Greece | - P. and A. Kyriakou Children’s Hospital, Athens | 9707/21.05.2020 |
| Italy | - Università degli Studi di Milano Statale, Milan - Ospedale Pediatrico Bambino Gesù, Rome | 0018932-U 27/05/202 & 848196 |
| Latvia | - Children’s Clinical University Hospital, Riga | Nr. 01-29.1/2736 |
| Netherlands | - University Medical Center Utrecht, Utrecht - Erasmus Medical Center., Rotterdam - Amsterdam University Medical Center, Amsterdam | 20-774/M (NL75190.041.20) |
| Slovenia | - University Medical Centre Ljubljana | 0120-271/2020/3 |
| Spain | - Hospital Clínico Universitario de Santiago de Compostela - Servicio Madrileño de Salud, Madrid | 2020/219 & 20/234 |
| Switzerland | - University Children’s Hospital, Universität Bern, Bern - University Children’s Hospital Zürich, Zürich | 2020-01556 |

**Supplementary Table 3.** Multiple linear regression of inverse rank normalised absorbance to *emm1* *S. pyogenes* cell wall extract relative to IVIG.

| Coefficient | b | 95% CI | P value |
| --- | --- | --- | --- |
| *Diagnostic category:*  Control or trivial  Unwell child* | –  -0.22 | –  -0.41 to -0.03 | 0.022 |
| *Recruitment site:*  Europe  UK | –  -0.01 | –  -0.19 to 0.17 | 0.9 |
| *Biological sex:*  Female  Male | –  0.07 | –  -0.10 to 0.25 | 0.4 |
| *Timing of sample at age 0-2 years:*  Before March 2020  After March 2020 | –  0.15 | –  -0.08 to 0.38 | –  0.2 |
| *Age irrespective of pandemic:*  0 months  6 months  1 year  2 years  *Age before March 2020:*  3 years  4 years  *Age after March 2020:*  3 years  4 years | 1.1  –  0.21  0.58  1.0  1.3  0.62  1.00 | 0.73 to 1.4  –  -0.09 to 0.50  0.27 to 0.89  0.66 to 1.4  0.95 to 1.7  0.21 to 1.0  0.62 to 1.4 | <0.001  0.17  <0.001  <0.001  <0.001  0.003  <0.001 |

***** Probable viral, unknown bacterial or viral, uncertain infection or inflammation or other unknown cause of illness

**Supplementary Table 4.** Multiple linear regression of inverse rank normalised absorbance to *emm12* *S. pyogenes* cell wall extract relative to IVIG.

| Coefficient | b | 95% CI | P value |
| --- | --- | --- | --- |
| *Diagnostic category:*  Control or trivial  Unwell child* | –  -0.28 | –  -0.47 to -0.09 | 0.004 |
| *Recruitment site:*  Europe  UK | –  -0.11 | –  -0.29 to 0.07 | 0.2 |
| *Biological sex:*  Female  Male | –  0.12 | –  -0.06 to 0.30 | 0.2 |
| *Timing of sample at age 0-2 years:*  Before March 2020  After March 2020 | –  0.17 | –  -0.06 to 0.39 | –  0.15 |
| *Age irrespective of pandemic:*  0 months  6 months  1 year  2 years  *Age before March 2020:*  3 years  4 years  *Age after March 2020:*  3 years  4 years | 1.1  –  0.24  0.62  0.98  1.4  0.63  1.1 | 0.73 to 1.4  –  -0.06 to 0.53  0.31 to 0.82  0.62 to 1.33  1.0 to 1.7  0.22 to 1.0  0.71 to 1.5 | <0.001  0.12  <0.001  <0.001  <0.001  0.003  <0.001 |

***** Probable viral, unknown bacterial or viral, uncertain infection or inflammation or other unknown cause of illness

**Supplementary Table 5.** Multiple linear regression of inverse rank normalised reactivity to RSV F-protein.

| Coefficient | b | 95% CI | P value |
| --- | --- | --- | --- |
| *Diagnostic category:*  Control or trivial  Unwell child* | –  0.17 | –  -0.02 to -0.36 | 0.077 |
| *Recruitment site:*  Europe  UK | –  0.02 | –  -0.17 to 0.21 | 0.8 |
| *Biological sex:*  Female  Male | –  -0.06 | –  -0.25 to 0.13 | 0.5 |
| *Timing of sample at age 0-2 & 4 years:*  Before March 2020  After March 2020 | –  -0.15 | –  -0.40 to 0.10 | –  0.2 |
| *Age irrespective of pandemic:*  0 months  6 months  1 year  2 years  4 years  *Age before March 2020:*  3 years  *Age after March 2020:*  3 years | 0.49  –  -0.24  0.28  1.2  1.3  0.52 | 0.11 to 0.87  –  -0.55 to 0.07  -0.03 to 0.58  0.85 to 1.5  0.95 to 1.7  0.02 to 1.0 | 0.011  0.13  0.080  <0.001  <0.001  0.040 |

***** Probable viral, unknown bacterial or viral, uncertain infection or inflammation or other unknown cause of illness

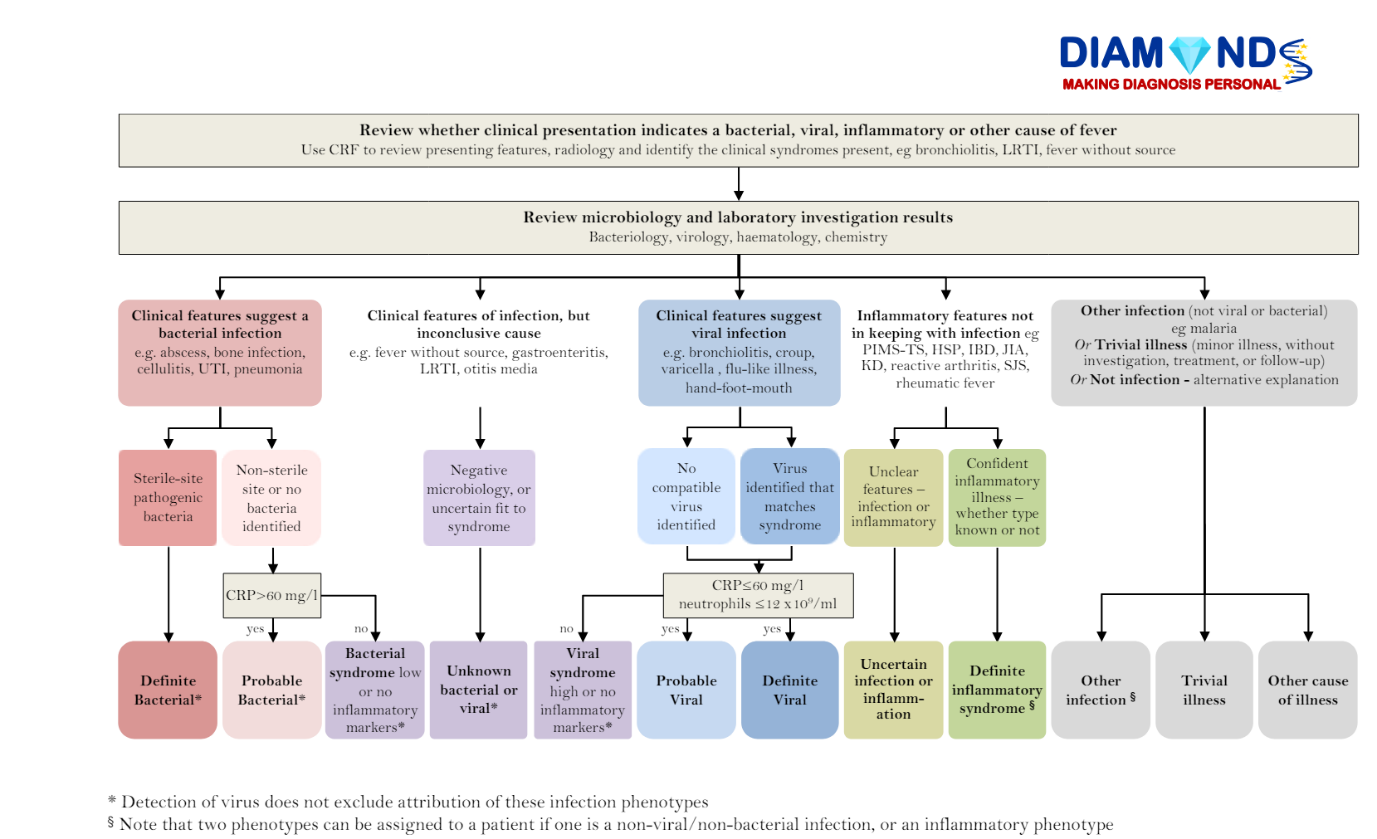

**Supplementary Figure 1.** Diagnostic algorithm used for classification.

PERFORM (N=491)

Controls – 241

Probable Viral – 226

Trivial Illness - 24

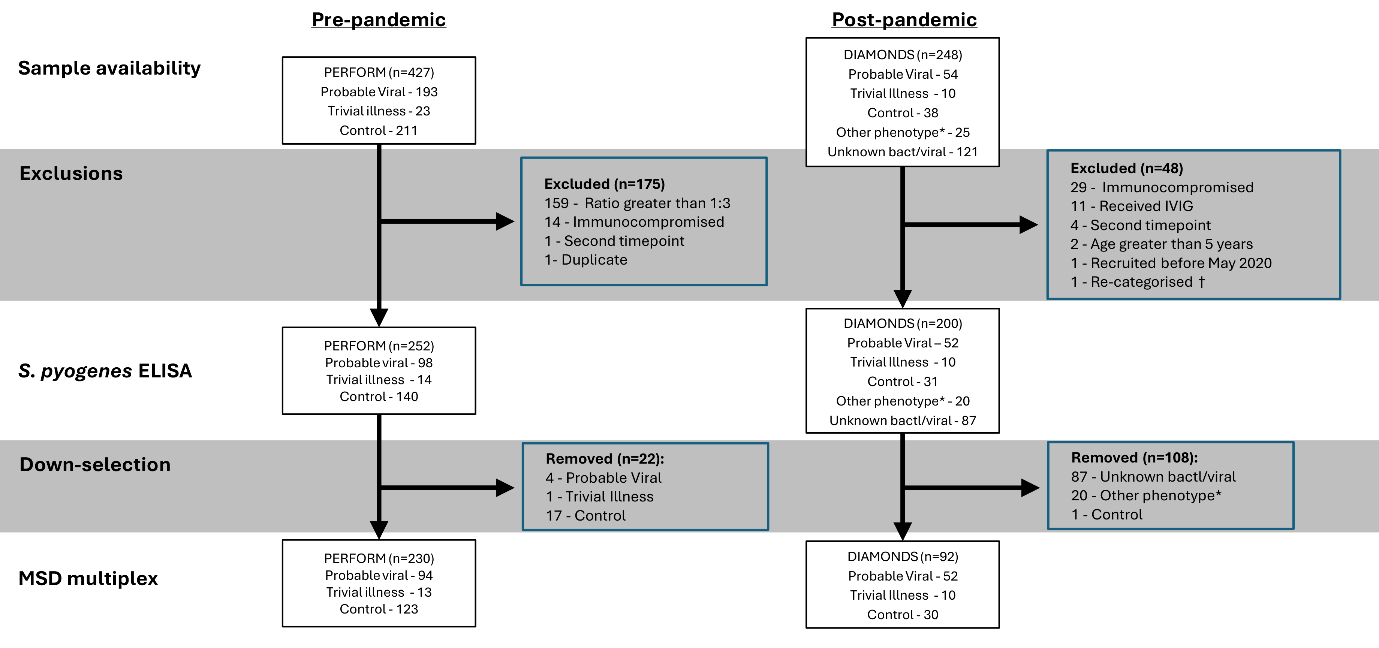

**Supplementary Figure 2.** Selection of samples for the antibody-mediated immunity study.

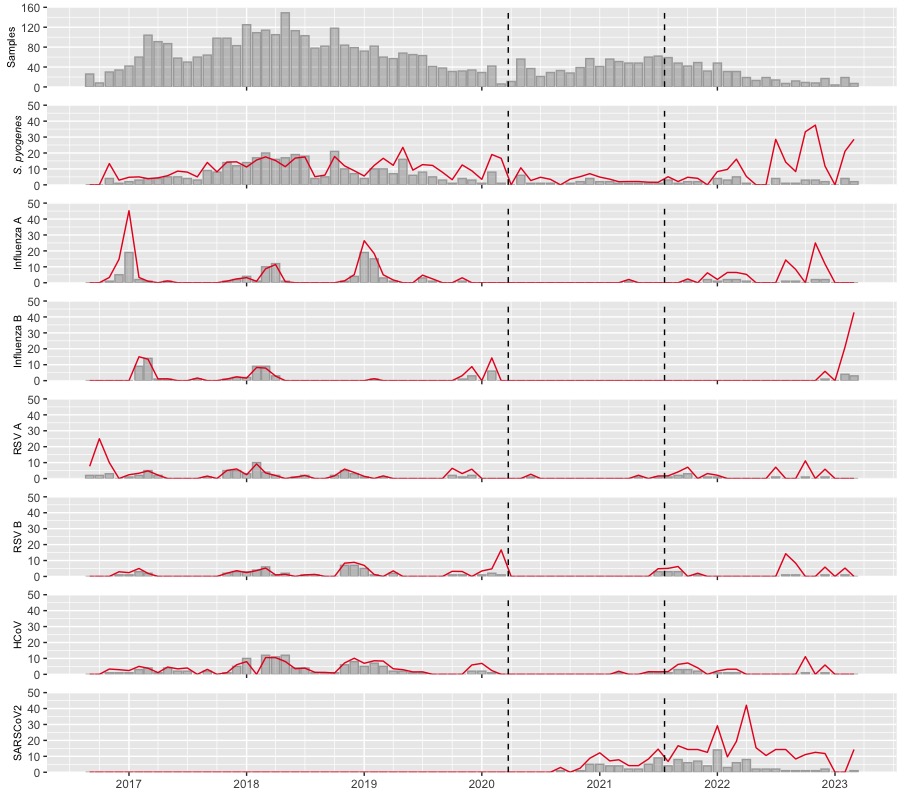

**Supplementary Figure 3.** Detection of *S. pyogenes* and viral pathogens by PCR among children aged 0-17 years (n=4,130) recruited to the PERFORM and DIAMONDS studies during September 2016 to August 2023. The monthly number of valid samples processed is shown in the first plot followed by the monthly number of positives for each pathogen indicated using grey bars. The red line indicates the monthly percentage of positive samples. Dashed vertical lines indicate the dates of introduction and cessation of restrictions in the UK. RSV, respiratory syncytial virus; HCoV, common cold coronaviruses 229, OC43, NL63 and HKU1 (combined); SARSCoV2, Severe acute respiratory syndrome coronavirus 2.

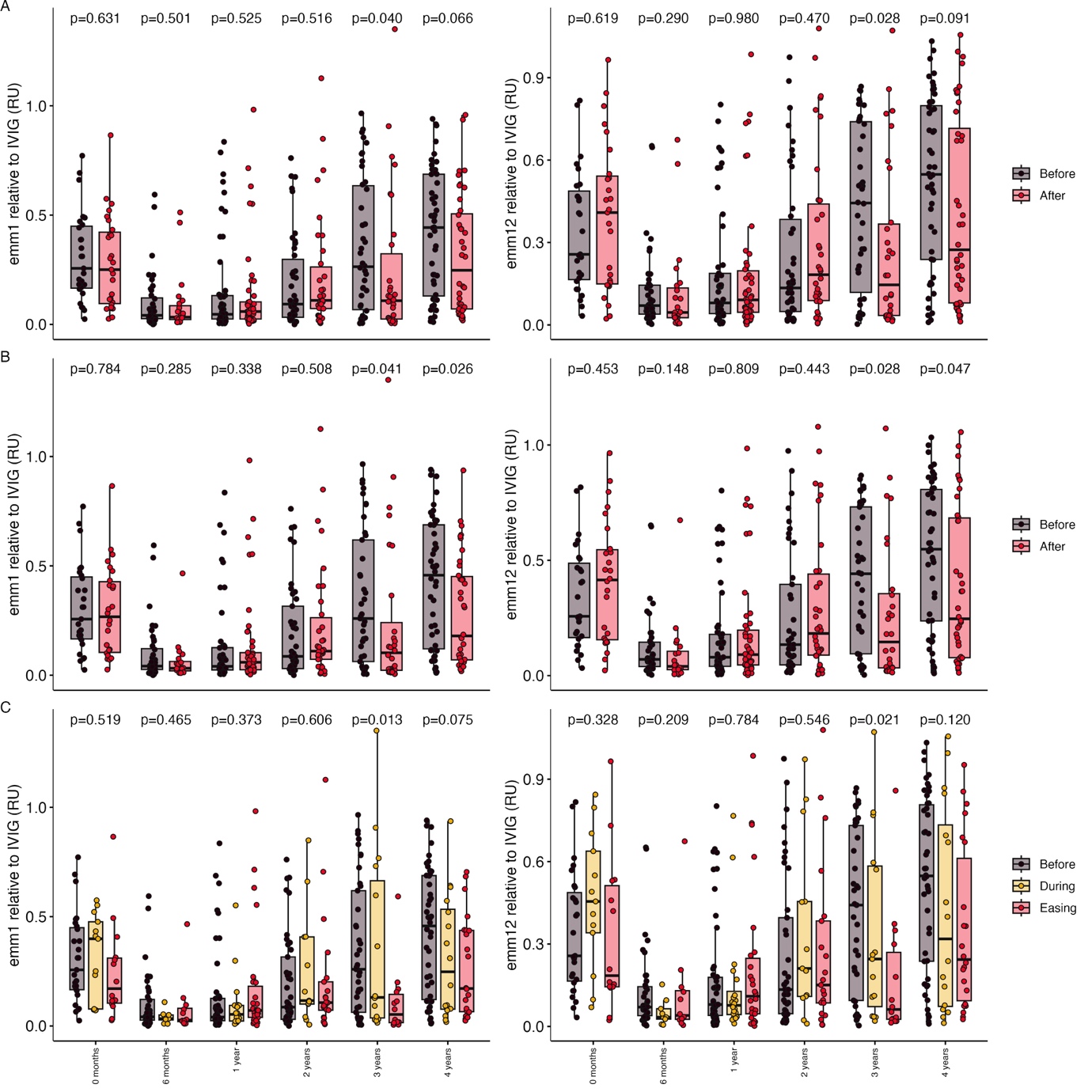
**Supplementary Figure 4. Reactivity to *S. pyogenes* by age band in sensitivity analyses.** Boxplots for *emm1* (left) and *emm12* (right) show absorption relative to IVIG: **A**. Irrespective of detection of *S. pyogenes* by PCR at the time of sampling; **B**. Excluding individuals with detection of SARS-CoV-2; and **C**. Dividing timing of recruitment into three periods. Differences between before (black) and after (red) March 2020 were assessed by a Wilcoxon rank-sum test, while differences before March 2020 (black), April 2020 to July 2021 (yellow) or after July 2021 (red) were assessed using a Kruskal-Wallis test.

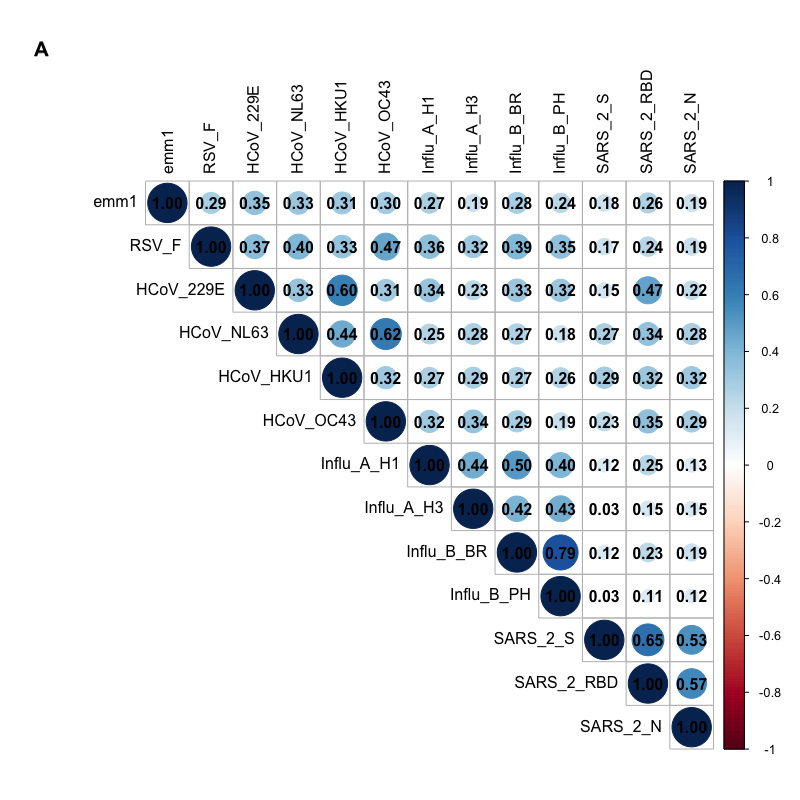

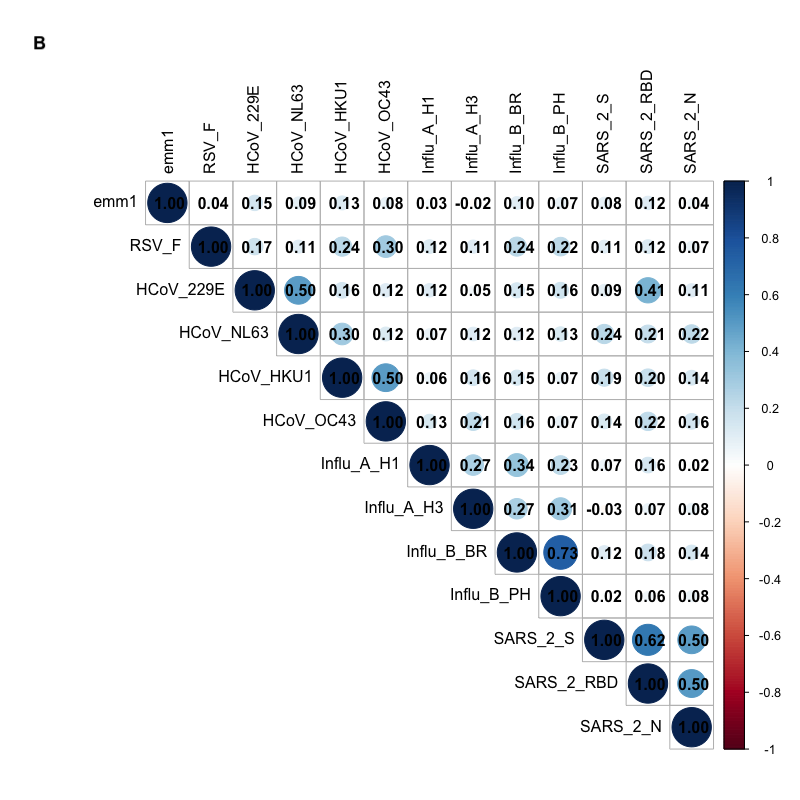

**Supplementary Figure 5.** Pearson’s correlation coefficient (*r*) for the relationship between reactivity to *S. pyogenes* and the 12 viral antigens in 230 samples used in the MSD assay. **A.** Unadjusted rank-normalised reactivity; **B.** Age-adjusted rank-normalised reactivity as the residuals from a linear regression model for rank-normalised reactivity with parameters for each of the age bands.

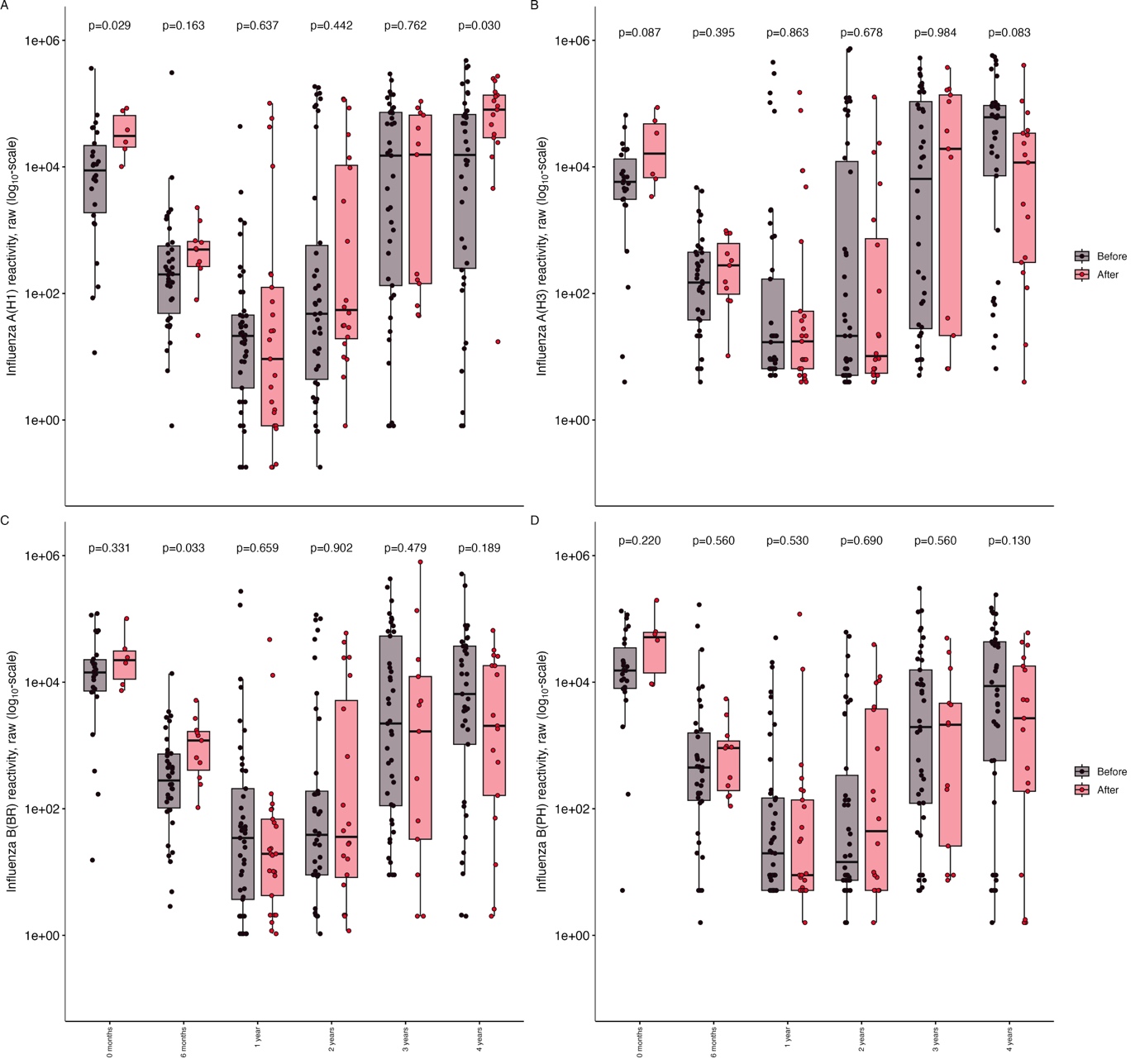
**Supplementary Figure 6. Reactivity to influenza virus haemagglutinins by age band.** Box plots show reactivity before (black) and after (red) March 2020 with the difference between groups assessed by a Wilcoxon rank-sum test. **A**. Influenza A (H1N1); **B.** Influenza A (H3N2); **C**. Influenza B (Brisbane); and **D**. Influenza B (Phuket).

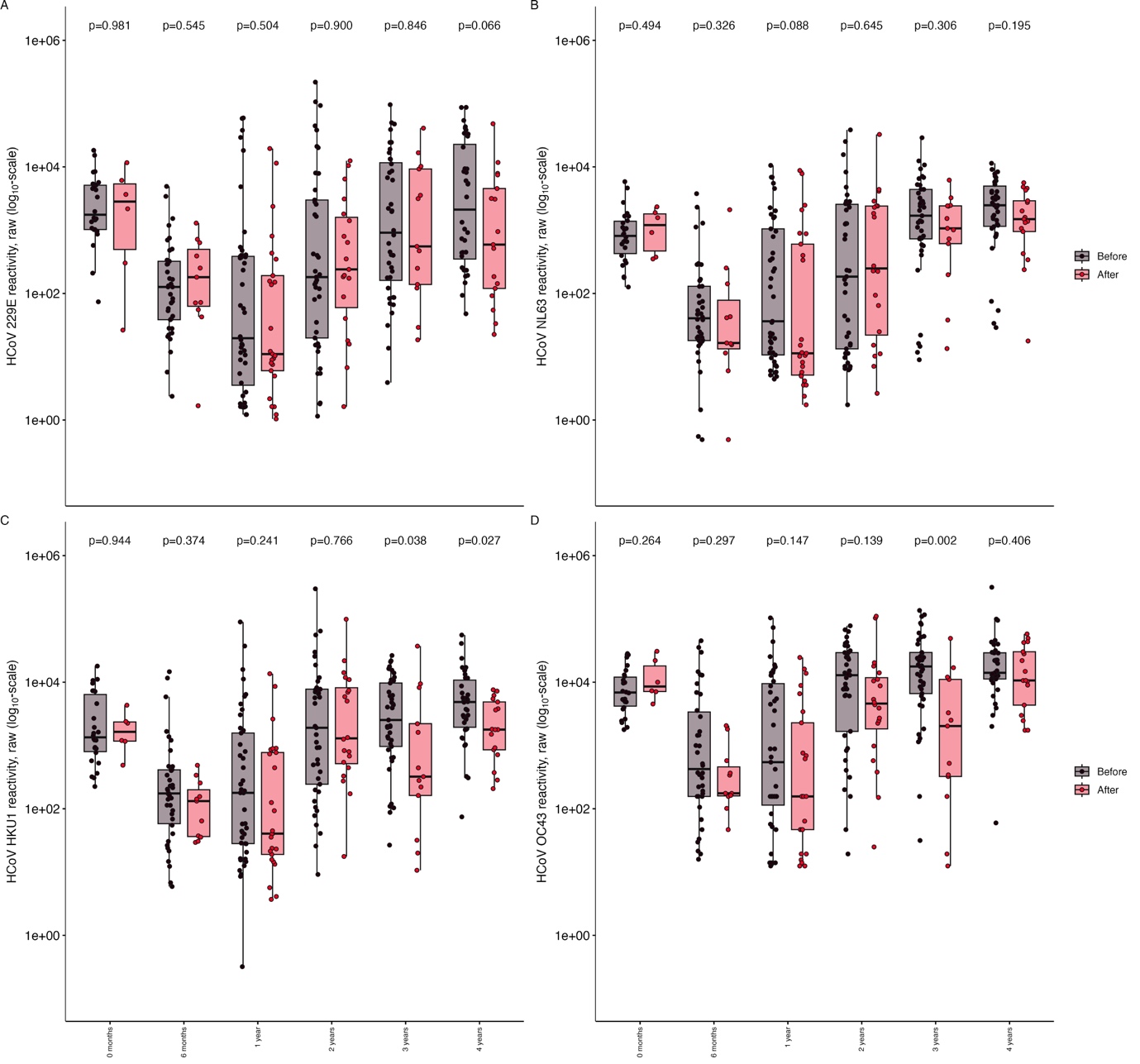
**Supplementary Figure 7. Reactivity to common cold coronavirus spike proteins by age band.** Box plots show reactivity before (black) and after (red) March 2020 with the difference between groups assessed by a Wilcoxon rank-sum test. **A**. 229E; **B.** HKU1; **C**. OC43; and **D**. NL63.

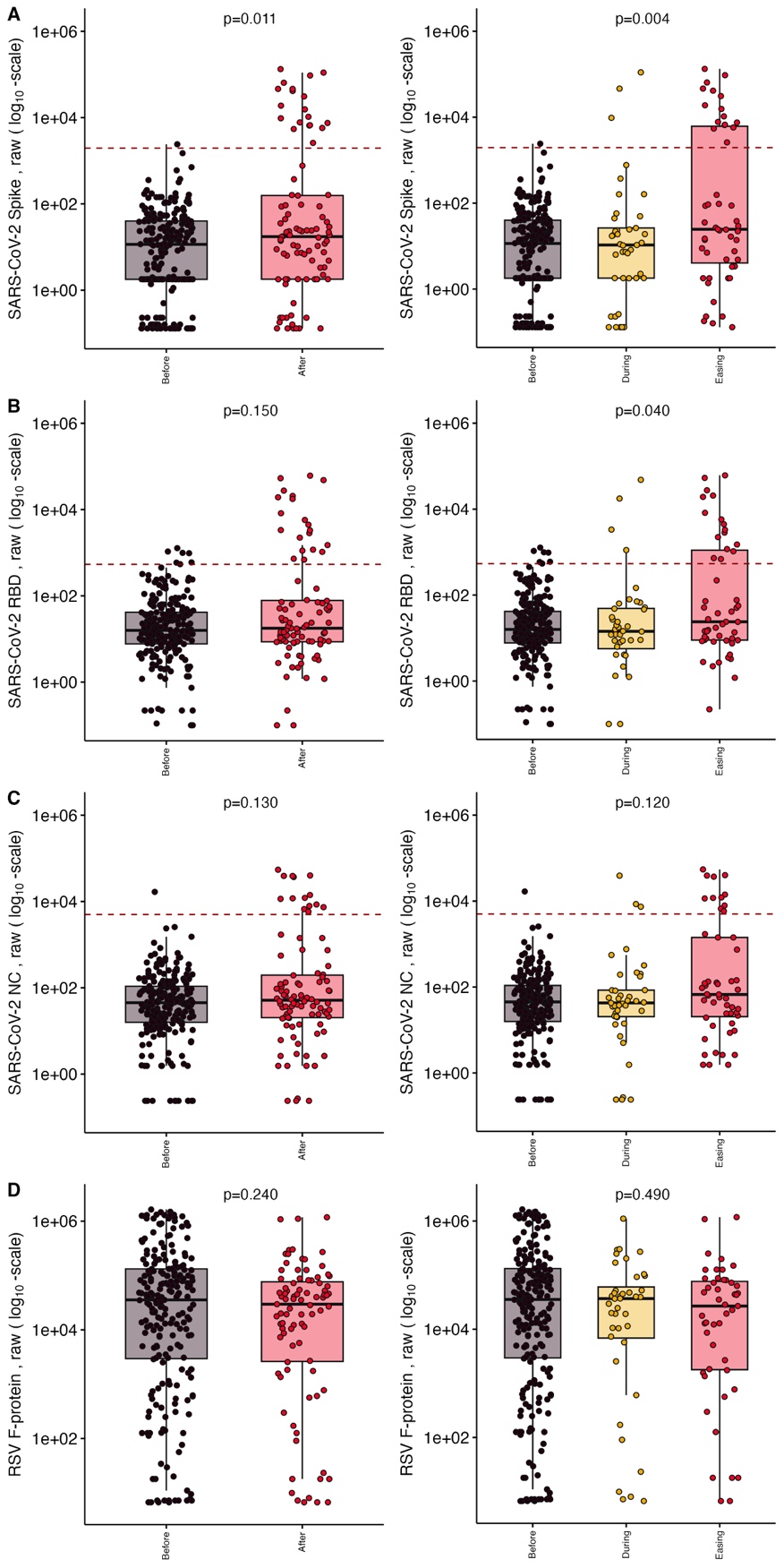

**Supplementary Figure 8.** **Reactivity to SARS-CoV-2 antigens and RSV F-protein across all ages.** Boxplots show reactivity between before (black) and after (red) March 2020 with differences assessed by a Wilcoxon rank-sum test (left), and before March 2020 (black), during April 2020 to July 2021 (yellow) or after July 2021 (red) with differences assessed using a Kruskal-Wallis test (right) for: **A**. SARS-CoV-2 Spike protein; **B.** SARS-CoV-2 Spike RBD; **C.** SARS-CoV-2 Nucleocapsid protein; and **D**. RSV F-protein, which was representative of distributions observed for other viruses. For the SARS-CoV-2 antigens, dashed horizontal lines indicate the manufacturer-recommend threshold for positivity.

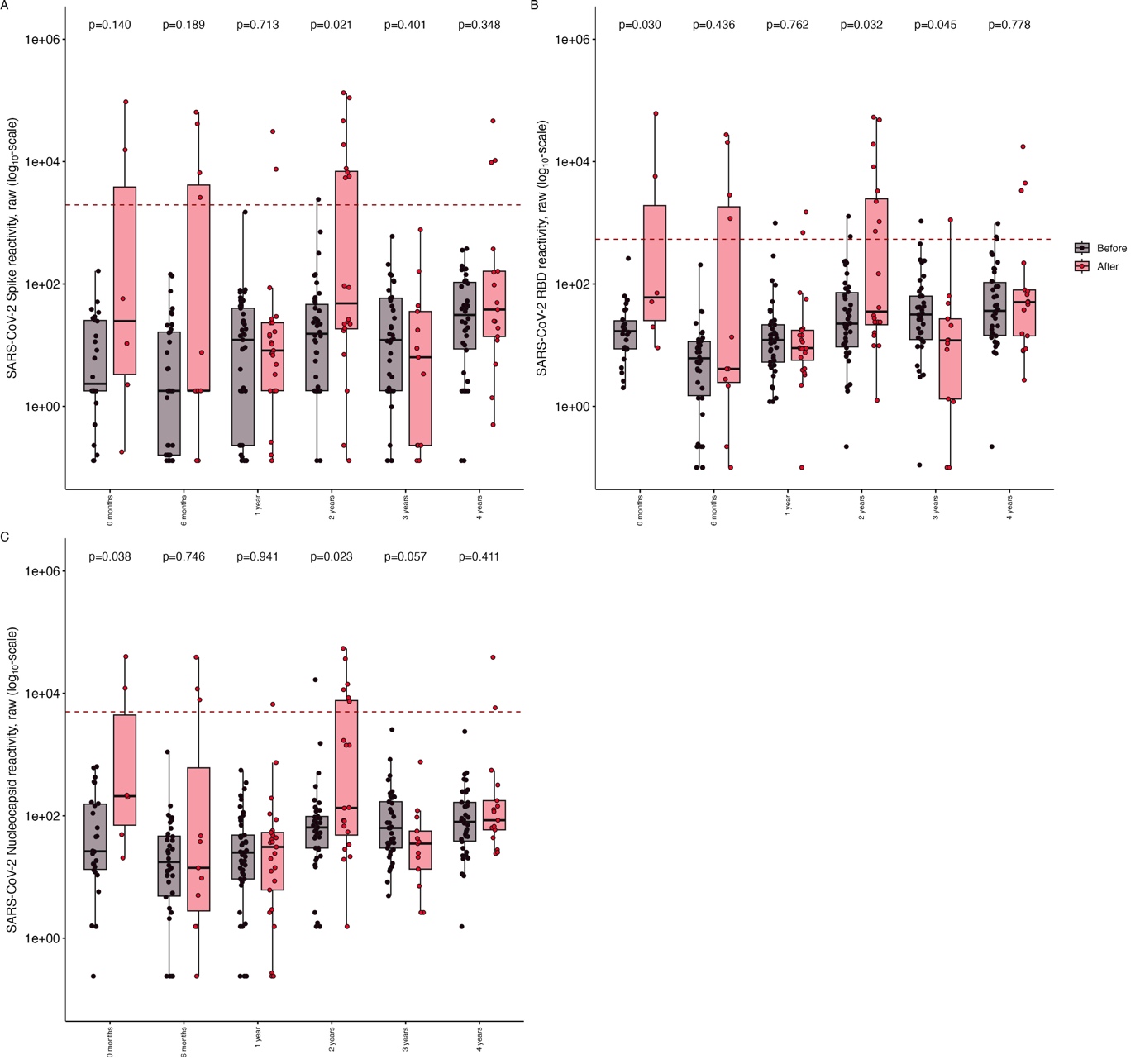
**Supplementary Figure 9. Reactivity to SARS-CoV-2 antigens by age band.** Box plots show reactivity before (black) and after (red) March 2020 with the difference between groups assessed by a Wilcoxon rank-sum test. **A**. SARS-CoV-2 Spike protein; **B.** SARS-CoV-2 Spike RBD; **C.** SARS-CoV-2 Nucleocapsid protein.

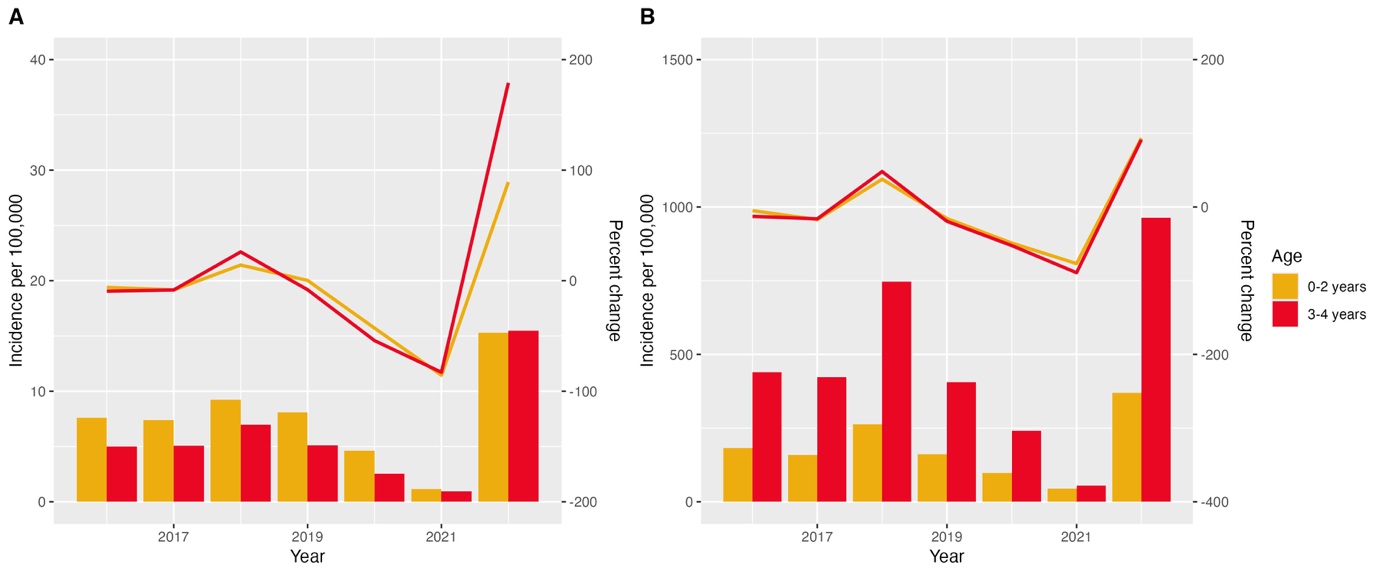
**Supplementary Figure 10. Changes in *S. pyogenes* disease incidence by age.** Bar charts showing incidence (left axis) per 100,000 person-years with a line indicating the percentage change (right axis) relative to 2016-2019. **A.** iGAS; **B.** Scarlet fever.
